# Accelerometer-derived Step Metrics and Quality of Life in Individuals with and without Cardiovascular Diseases

**DOI:** 10.64898/2026.07.30.26359231

**Authors:** Sabine Schootemeijer, Sophie H. Kroesen, Niels A. Stens, Milou Netten, Inge P. Salzmann, Annemarie Koster, Neeltje A.E. Allard, Bram M.A. van Bakel, Francisco B. Ortega, Emmanuel Stamatakis, Matthew Ahmadi, Janna N. Vrijsen, Dick Thijssen, Thijs M.H. Eijsvogels, Esmée A. Bakker, the STEP COACH collaborators

## Abstract

**Background and Aims:** Steps are increasingly used to prescribe physical activity, but their impact on quality of life (QoL) remains unclear. We examined the dose-response association between step metrics and QoL, and whether cardiovascular disease (CVD) status moderates this association.

**Methods:** Individual-level data of five studies were pooled. Physical activity was measured with thigh-worn accelerometry. We assessed steps/day, daily minutes of fast stepping (≥100 steps/min), peak 1- and 30- min cadence. We investigated the association of step metrics and QoL (questionnaire-based; standardized) with multivariable (non-)linear regression, and the interaction with CVD status.

**Results:** We included 9,371 participants (62 [54-68] years; 47% female), comprising 1,977 individuals with and 7,394 without CVD. Significant, curvilinear dose-response associations between step metrics and QoL were found. The optimal step volume was 6,561 steps/day which associated with a 0.35 SD (95%CI: 0.28-0.42) higher QoL compared to the referent 4,000 steps/day. The optimal doses for peak 1-min and peak 30-min cadence were 107 steps/minute (+0.34 SD; 95%CI: 0.28-0.41) and 74 steps/minute (+0.29 SD; 95%CI: 0.24-0.34) respectively, compared to references of 90 and 60 steps/minute. Only fast stepping interacted with CVD status, with a lower optimum in those with *versus* without CVD (4 minutes/day, +0.20 SD, 95%CI: 0.12-0.28 *versus* 9 minutes/day, +0.19 SD, 95%CI: 0.12-0.25), compared to the referent 2 minutes/day.

**Conclusions:** Step metrics were curvilinearly associated with QoL with optimal benefits at ∼6,500 steps/day. Optimal QoL benefits can be reached at feasible stepping targets, and at slightly fewer daily minutes of fast stepping in CVD versus non-CVD.

**Structured graphical abstract:** *Key Questions:*

- Is there a dose-response association between step metrics and health-related quality of life (QoL)?
- Does the cardiovascular disease (CVD) health status moderate this association?

*Key Findings:*

- More steps/day and time spent at fast stepping are associated with better QoL following a curvilinear dose-response curve.
- The optimal step volume is 6,561 steps/day, which was independent of CVD status.
- Individuals with CVD may gain optimal QoL levels at slightly less time of fast stepping (4 min/day) than those without CVD (9 min/day).

**Take Home Message:** More steps and steps at a higher intensity are associated with better QoL. Optimal QoL benefits could be gained at feasible stepping targets (i.e. 4 min/day of fast stepping for CVD, 9 min/day for those without CVD).

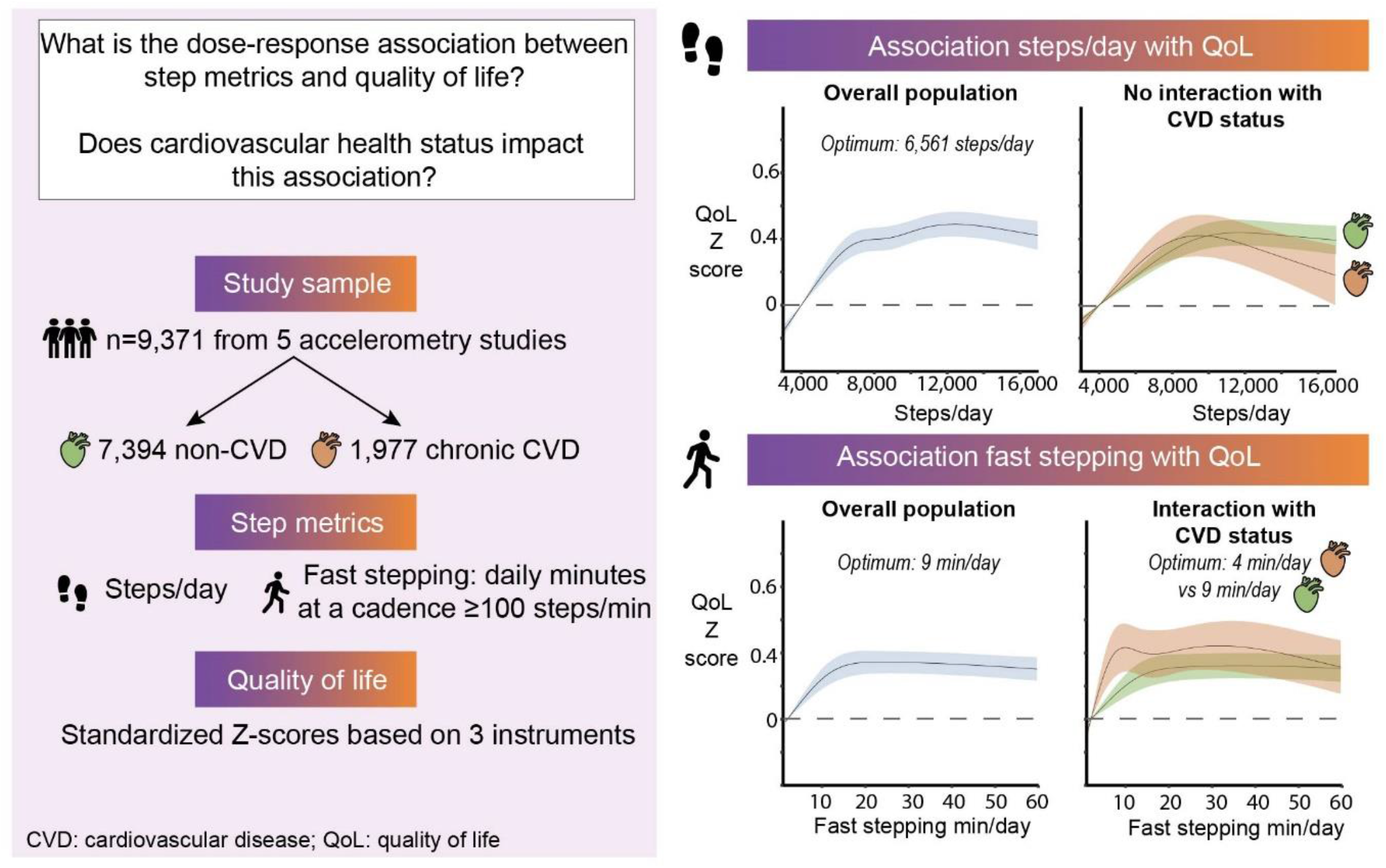

## Introduction

A physically active lifestyle improves health and lowers the risk of chronic diseases.[1] Moreover, higher volumes of physical activity (PA) are associated with a better health-related quality of life (QoL) in healthy adults[2-5] and individuals with cardiovascular disease (CVD).[6-8] However, most evidence is based on self-reported PA data [3-7] which is prone to recall bias (i.e., over- or underreporting PA) and does not capture habitual, unstructured PA or step metrics. Wearable-derived step metrics, i.e. measures of step volume and step intensity, may provide an easy-to-understand, objective measure of PA. Insight into the association with QoL could help individuals to improve their mental and physical health.

Emerging evidence suggests that the dose-response associations between PA and health outcomes are dependent on health status.[9, 10] For example, a curvilinear association between moderate-to-vigorous PA and adverse cardiovascular events and mortality was found in healthy individuals, whereas a linear association was present in individuals with CVD.[10] Delineating whether the dose-response relationship between daily step metrics and QoL is different between individuals with versus without CVD, is key for personalized PA prescription. Step metrics could for instance be implemented in wearable-based cardiac rehabilitation programs to increase physical activity levels.[11, 12]

Therefore, we sought to examine the cross-sectional dose-response association between step metrics and QoL, and to delineate whether this association differs between those with and without CVD. We hypothesize that step metrics are positively associated with QoL and that the shape of the dose-response curve differs between those with (linear) and without CVD (curvilinear).

## Methods

### Study design

We pooled cross-sectional data from individuals with and without CVD from three observational cohort studies (i.e. The Maastricht Study (TMS; n=7,704)[13], Nijmegen Exercise Study (NES; n=2,007)[14]) and The Physical Activity in Statin Users Study (FACTS; n=194)[15]) and two randomized controlled trials (i.e., Cardiac RehApp, n=37[16] and SIT LESS, n=178[17]). Details of the included studies are provided in **Supplementary Table 1**. All cohorts recruited participants from the general population including those with chronic CVD, whereas the randomized controlled trials performed prospective measurements in CVD patients during and following cardiac rehabilitation. As QoL improves during the rehabilitation phase after a cardiovascular event [18, 19], we included the final assessment of daily steps from the randomized controlled trials, conducted at 3 months following graduation of the cardiac rehabilitation program. In this way, the assessment of steps is as close to the chronic phase as possible [16, 17]. Inclusion criteria for our data analyses were the availability of i) a completed QoL questionnaire and ii) a valid device-based PA assessment using thigh-worn accelerometry. The requirement of ethical approval for harmonization and additional data analysis was waived by the ethics committee of the Radboud university medical center, Nijmegen, The Netherlands (#2025-18512).

### Study sample

Participants with a diagnosis of coronary artery disease (myocardial infarction, angina pectoris with the use of CVD medication, previous percutaneous coronary intervention or other surgery on heart vessels), heart failure, valvular heart disease, arrhythmias (including atrial fibrillation), cerebral vascular event and peripheral artery disease were classified as the CVD group. CVD diagnoses were retrieved from medical records or based on self-report, dependent on the study (**Supplementary Table 1**). All participants without overt CVD were classified as ‘non-CVD’.

### Quality of life

QoL was assessed with three different questionnaires. FACTS and NES included a one-item question about overall QoL (“How would you grade your quality of life” (1=very poor, 10=excellent)). Cardiac RehApp and SITLESS used a disease-specific QoL questionnaire, the HeartQoL.[20, 21] TMS used the Short-Form-36 version 1 (SF-36)[22], for which we used the SF6D-utility index[23, 24] to estimate overall QoL. Furthermore, we calculated physical and mental component scores for the studies that applied the SF-36 and the HeartQoL.[20-22, 25, 26] For all instruments, higher scores indicated better QoL. To analyze the data from different QoL instruments, we standardized the QoL scores per questionnaire meaning that we combined studies using the same questionnaire and calculated a pooled standard deviation (SD), as suggested by the NICE guidelines.[27] Using this approach the SD is weighted for the sample size of the individual studies. Z-scores were calculated by subtracting the mean scores from the individual scores divided by the pooled SD. The QoL z-scores correspond to Cohen’s d effect sizes (negligible effect: ≤0.2, small effect: 0.2-0.5, medium effect: 0.5-0.8, large effect: ≥0.8).[28]

### Step metrics

PA was assessed with a thigh-worn accelerometer (activPAL 3 micro, PAL Technologies, Glasgow, UK) and participants were instructed to wear the accelerometer for eight days and 24 hours/day. Raw accelerometry data were processed with the validated ActiPASS algorithm (v2024.06.01).[29] A day was considered valid if the accelerometer was worn for ≥20 hours, had ≥1 walking bout, >0 min of sleep, and ≥1,000 steps/day. A measurement was included when there were at least four valid days including one weekend day. For all valid measurements, the ActiPASS output files containing steps per minute were processed by the STRIDE script (v0.2.2) in Python (v3.13) to obtain steps metrics. The average step volume per day was calculated and used as our main exposure. Additionally, three steps metrics representing step intensity were determined; 1) fast stepping (minutes/day at a cadence ≥100 steps/minute[30]), 2) peak 1-minute cadence (steps/min), and 3) peak 30-minute cadence (steps/min).[31] Peak 1-minute cadence was calculated by averaging the cadence during the minute with the highest cadence of each valid measurement day. Peak 30-minute cadence was calculated by averaging the cadence during the 30 minutes with the highest cadence of each day.[31] Steps and cadence metrics were only considered during activities classified by ActiPASS as moving (i.e., standing that involves movement), walking, stair climbing and running.

### Covariates

We adjusted the analyses for the following, harmonized, covariates: age (years), sex (male/female), BMI (kg/m^2^), alcohol use (current alcohol use yes/no), smoking status (current smoker yes/no) and diabetes mellitus (yes/no). Details about the harmonization of covariates across cohorts are presented in **Supplementary Table 2**.

### Statistical analyses

Descriptive data were presented as mean (SD) or median (interquartile range; Q25-Q75) depending on the normal distribution, and as number (%) for categorical data. After data processing and harmonization, we analyzed individual participant data with multivariable regression models. Since <10% of data on the covariates were missing, we conducted a complete cases analysis. To assess the presence of a non-linear association, we applied restricted cubic spline regression models. We tested 3 knots (located at 10^th^, 50^th^, and 90^th^ centiles), 4 knots (at 5^th^, 35^th^, 65^th^, and 95^th^ centiles), and 5 knots (at 5^th^, 27.5^th^, 50^th^, 72.5^th^, and 95^th^ centiles) against the referent values, and determined the best model fit based on statistical significance (p<.05) of the likelihood ratio test. If the non-linear model showed the best fit, non-linearity was verified with the Wald statistic. Model assumptions, including the normality of the residuals, were verified. Reference values were set at the ±5^th^ percentile, corresponding to 4,000 steps/day for step count, 2 minutes for fast stepping, 90 steps/minute for peak 1-min cadence and 60 steps/minute for peak 30-min cadence. We defined the optimum dose for each step metric as the highest QoL that was achieved at the least effort.[32] This corresponds to the lowest step metric value at which the upper limit of the 95% CI exceeded the lower bound of the 95% CI at higher step values and QoL scores (**Supplementary Figure 1**). For our main aim, we also report the dose with the maximum effect, defined as the step volume corresponding to the highest estimated QoL.

To investigate whether the associations between steps metrics and QoL differed between individuals with and without CVD, we tested for interaction with CVD health status and conducted stratified analyses. We tested the interaction term between each step metric and CVD status to estimate whether the dose-response association was dependent on CVD status. To evaluate the robustness of our results, we 1) examined the dose-response association using five categories for step metrics (e.g. for step volume: ≤5,000 (reference), 5,000-7,500, 7,500-10,000, 10,000-12,500, >12,500 steps/day) and stratified our main analysis for 2) QoL instrument and 3) cohort. Details of other sensitivity analyses that tested for effect modification of age, sex, BMI and CVD type, and an analysis excluding individuals with other chronic diseases are described in the statistical analyses plan (SAP).[33]

Statistical analyses were conducted in Rstudio version 2024.4.2.764 [34] using ‘rms’ for the restricted cubic splines. All statistical tests were two-tailed and p-values for statistically significance were set at 0.05. The SAP was pre-registered prior to the analyses and updated when needed, and is available at the Open Science Framework.[33]

## Results

### Study population

From the 10,120 individuals eligible for inclusion, 9,371 (93%) were included in our analyses, of which 1,977 (21%) individuals reported a diagnosis of CVD (**Supplementary Figure 2**). Participants with CVD were older (66 [60–71] *versus* 61 [53-67] years), had a higher BMI (27.0 [24.5–29.8] *versus* 25.5 [23.2– 28.4]) and were less often employed than those without CVD (27% *versus* 47%) (**Table 1**). Individuals with CVD took fewer steps/day, spent less time at fast stepping cadence, and had a lower peak 1-min and peak 30-min cadence than those without CVD (**Table 1; Supplementary Figure 3**). Instrument-specific overall QoL scores were high in our study population (HeartQoL (range 0-3): 2.0 [1.6–2.8]; SF6D (range 0-1): 0.8 [0.8–0.9]; 1-item (range 1-10): 8.0 [8.0–9.0]; **Supplementary Table 3**).

**Table 1.**
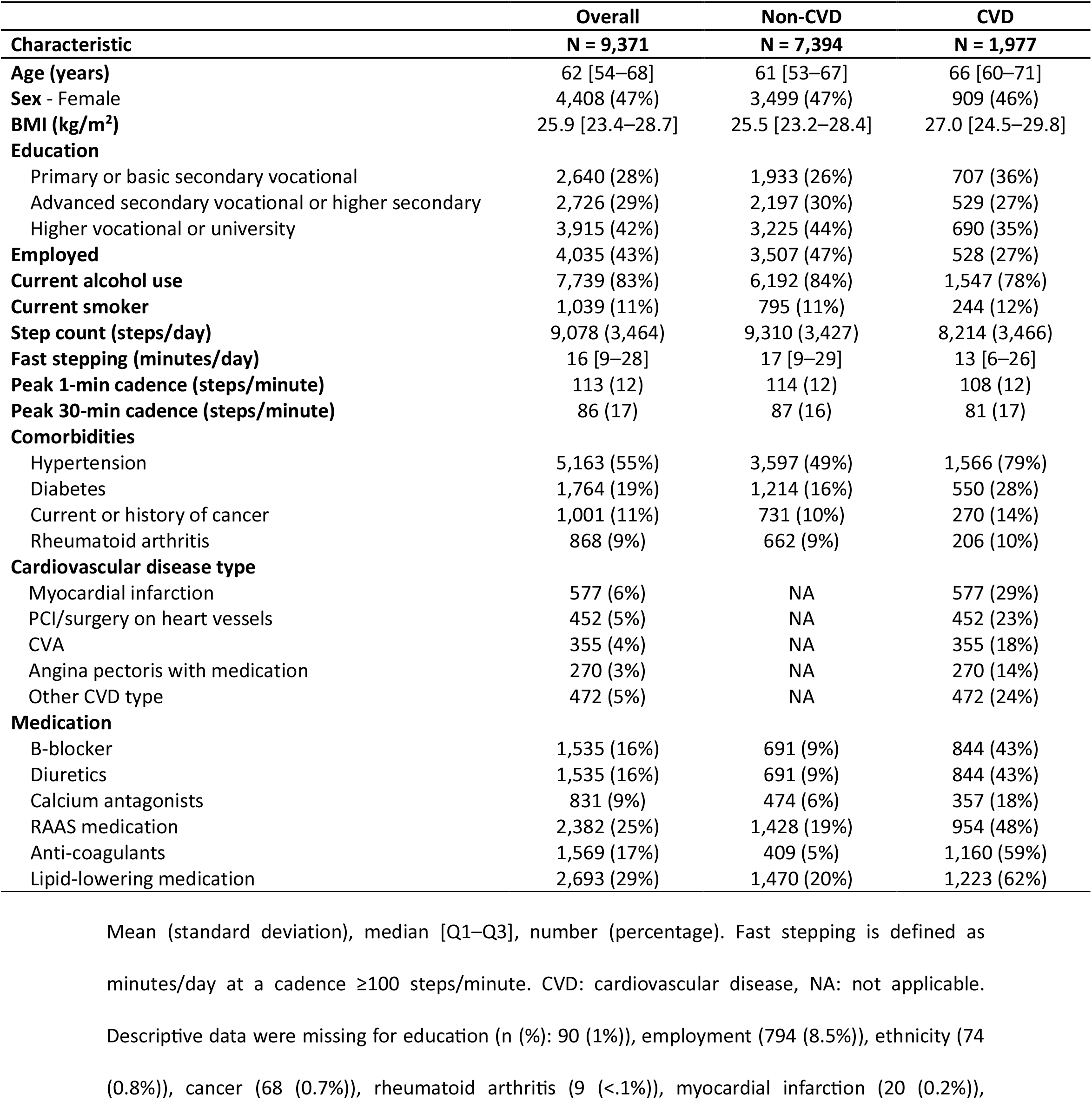
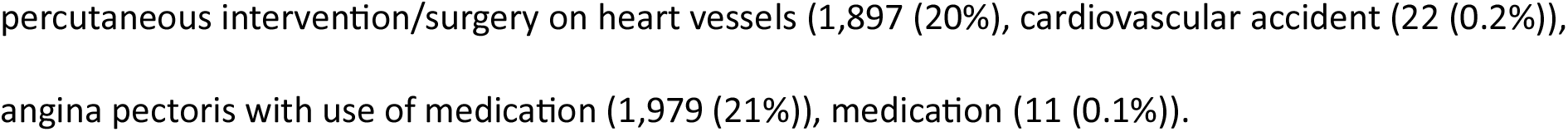
Descriptive characteristics of the overall study sample and stratified by cardiovascular health status.

| Characteristic | Overall<br>N = 9,371 | Non-CVD<br>N = 7,394 | CVD<br>N = 1,977 |
| --- | --- | --- | --- |
| **Age (years)** | 62 [54–68] | 61 [53–67] | 66 [60–71] |
| **Sex - Female** | 4,408 (47%) | 3,499 (47%) | 909 (46%) |
| **BMI (kg/m<sup>2</sup>)** | 25.9 [23.4–28.7] | 25.5 [23.2–28.4] | 27.0 [24.5–29.8] |
| **Education** |  |  |  |
| Primary or basic secondary vocational | 2,640 (28%) | 1,933 (26%) | 707 (36%) |
| Advanced secondary vocational or higher secondary | 2,726 (29%) | 2,197 (30%) | 529 (27%) |
| Higher vocational or university | 3,915 (42%) | 3,225 (44%) | 690 (35%) |
| **Employed** | 4,035 (43%) | 3,507 (47%) | 528 (27%) |
| **Current alcohol use** | 7,739 (83%) | 6,192 (84%) | 1,547 (78%) |
| **Current smoker** | 1,039 (11%) | 795 (11%) | 244 (12%) |
| **Step count (steps/day)** | 9,078 (3,464) | 9,310 (3,427) | 8,214 (3,466) |
| **Fast stepping (minutes/day)** | 16 [9–28] | 17 [9–29] | 13 [6–26] |
| **Peak 1-min cadence (steps/minute)** | 113 (12) | 114 (12) | 108 (12) |
| **Peak 30-min cadence (steps/minute)** | 86 (17) | 87 (16) | 81 (17) |
| **Comorbidities** |  |  |  |
| Hypertension | 5,163 (55%) | 3,597 (49%) | 1,566 (79%) |
| Diabetes | 1,764 (19%) | 1,214 (16%) | 550 (28%) |
| Current or history of cancer | 1,001 (11%) | 731 (10%) | 270 (14%) |
| Rheumatoid arthritis | 868 (9%) | 662 (9%) | 206 (10%) |
| **Cardiovascular disease type** |  |  |  |
| Myocardial infarction | 577 (6%) | NA | 577 (29%) |
| PCI/surgery on heart vessels | 452 (5%) | NA | 452 (23%) |
| CVA | 355 (4%) | NA | 355 (18%) |
| Angina pectoris with medication | 270 (3%) | NA | 270 (14%) |
| Other CVD type | 472 (5%) | NA | 472 (24%) |
| **Medication** |  |  |  |
| B-blocker | 1,535 (16%) | 691 (9%) | 844 (43%) |
| Diuretics | 1,535 (16%) | 691 (9%) | 844 (43%) |
| Calcium antagonists | 831 (9%) | 474 (6%) | 357 (18%) |
| RAAS medication | 2,382 (25%) | 1,428 (19%) | 954 (48%) |
| Anti-coagulants | 1,569 (17%) | 409 (5%) | 1,160 (59%) |
| Lipid-lowering medication | 2,693 (29%) | 1,470 (20%) | 1,223 (62%) |

4 Mean (standard deviation), median [Q1–Q3], number (percentage). Fast stepping is defined as 5 minutes/day at a cadence $\geq 100$ steps/minute. CVD: cardiovascular disease, NA: not applicable. 6 Descriptive data were missing for education (n (%): 90 (1%)), employment (794 (8.5%)), ethnicity (74 7 (0.8%)), cancer (68 (0.7%)), rheumatoid arthritis (9 (<.1%)), myocardial infarction (20 (0.2%)),
- 1 percutaneous intervention/surgery on heart vessels (1,897 (20%), cardiovascular accident (22 (0.2%)), - 2 angina pectoris with use of medication (1,979 (21%)), medication (11 (0.1%)).

### Dose-response association between step metrics and QoL

Step volume was positively associated with overall-, physical- and mental QoL following a curvilinear association (p for non-linearity <.05; **Figure 1**). The optimum number of daily steps was found at 6,561 steps/day and associated with a 0.35 SD (95%CI: 0.28-0.42) higher overall QoL compared to the referent 4,000 steps/day (**Figure 1A**). The maximum effect was observed at much higher steps: 12,427 steps/day were associated with a 0.49 SD (95%CI: 0.41-0.56) higher overall QoL. For physical QoL, we observed a greater effect size than for overall QoL, as the optimum of 8,274 steps/day was associated with a 0.44 SD (95%CI: 0.37-0.51) higher physical QoL (**Figure 1B**). The maximum effect was at 13,218 steps/day (+0.59 SD physical QoL, 95%CI: 0.51-0.66). Higher daily step volumes were associated with a higher mental QoL, but with a negligible effect size. The optimum dose was 5,516 steps/day (+0.05 SD, 95%CI: 0.02-0.08; **Figure 1C**) and the maximal dose 20,350 steps/day (+0.16 SD, 95%CI: 0.02-0.30).

**Figure 1.**
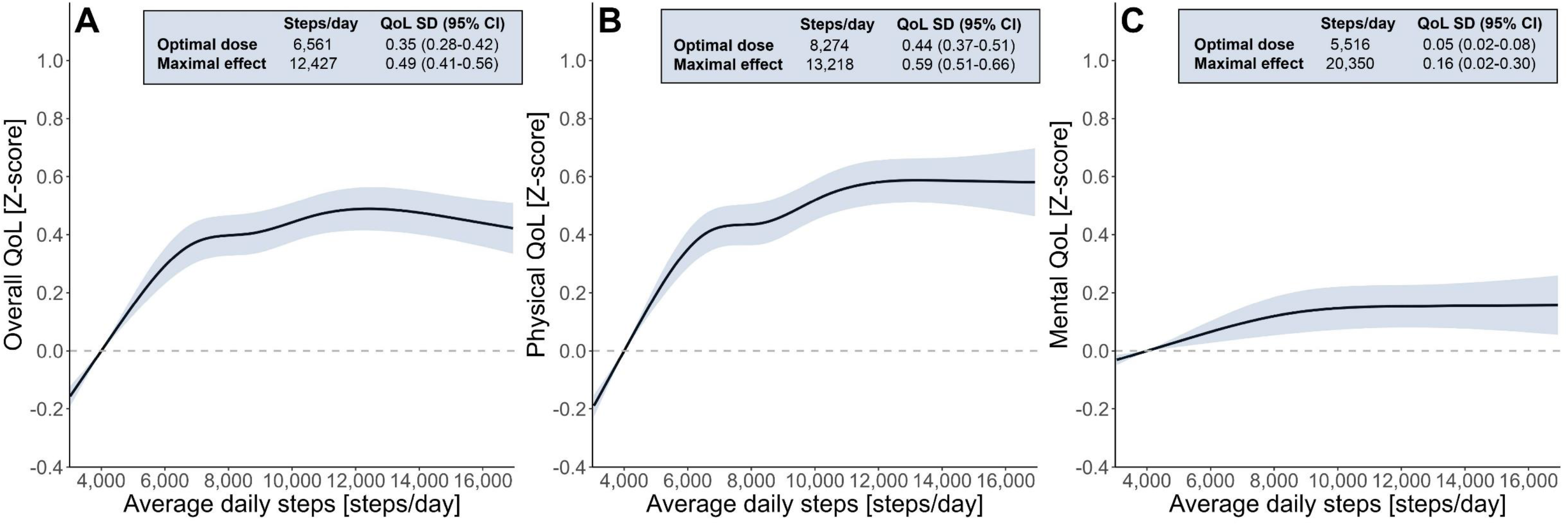
Dose-response association between steps/day and overall (A), physical (B) and mental (C) quality of life for the total population. Reference values are set at 4,000 steps/day. Shading represents the 95% confidence interval. The model was adjusted for age, sex, body mass index, alcohol consumption, smoking status and diabetes. QoL: quality of life.

Step intensity metrics followed a curvilinear dose-response curve with overall- and physical QoL (p for non-linearity <.05; **Figure 2A-F**), whereas linear dose-response curves were found for mental QoL (**Figure 2G-I**). The optimum of fast stepping was achieved at 9 minutes/day (+0.21 SD overall QoL, 95%CI: 0.16-0.27) compared to the referent 2 minutes/day (**Figure 2A**). Analyses of the physical component revealed a similar optimum but a greater effect size for physical (10 minutes/day, +0.46 SD overall QoL, 95%CI: 0.38-0.54) *versus* overall QoL, whereas no association with mental QoL was found (**Figure 2G**). A higher peak 1- and peak 30-min cadence was associated with a better overall-, physical- and mental QoL (**Figure 2**). Optimum cadence effects on overall QoL were found at 107 steps/minute for peak 1-min cadence (+0.34 SD, 95%CI: 0.28-0.41) and 74 steps/minute for peak 30-min cadence (+0.29 SD, 95%CI: 0.24-0.34), compared to reference values of 90 and 60 steps/minute, respectively. Our categorical regression analyses including five subgroups of step metrics were consistent with our main findings derived from the restricted cubic splines (**Table 2, Supplementary Table 4-5, Supplementary Figure 4-7**).

**Figure 2.**
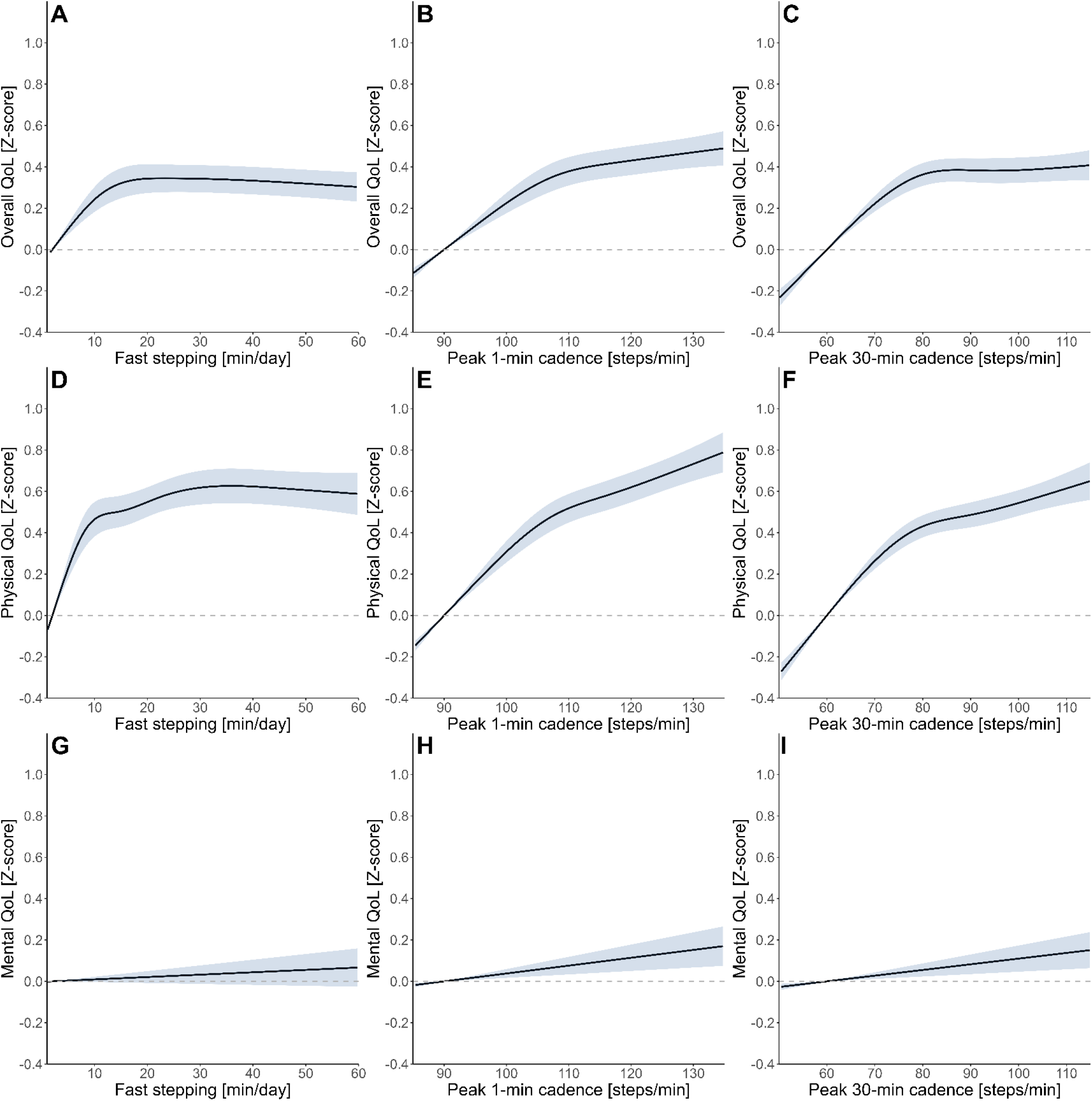
Dose-response association fast stepping (A,D,G), peak 1-min cadence (B,E,H) and peak 30- min cadence (C,F,I) and overall, mental and physical quality of life for the total population. Reference values are set at 2 minutes fast stepping, 90 minutes for peak 1-min cadence, 60 minutes for peak 30- min cadence. Shading represents the 95% confidence interval. The model was adjusted for age, sex, body mass index, alcohol consumption, smoking status and diabetes. QoL: quality of life.

**Table 2.** The associations between categories of step metrics with overall quality of life for the total population and stratified by cardiovascular disease status.

|  |  | Total population |  | Non-CVD |  | CVD |  |
| --- | --- | --- | --- | --- | --- | --- | --- |
| Step metric | n/CVD | Estimate (95% CI) | p-value | Estimate (95% CI) | p-value | Estimate (95% CI) | p-value |
| **Steps/day** |  |  |  |  |  |  |  |
| ≤5,000 | 876/340 | 1 |  | 1 |  | 1 |  |
| 5,000-7,500 | 2,421/588 | 0.30 (0.23, 0.38) | <.001 | 0.28 (0.18, 0.37) | <.001 | 0.34 (0.20, 0.48) | <.001 |
| 7,500-10,000 | 2,825/537 | 0.40 (0.33, 0.48) | <.001 | 0.40 (0.31, 0.49) | <.001 | 0.39 (0.24, 0.53) | <.001 |
| 10,000-12,500 | 1,917/308 | 0.47 (0.39, 0.56) | <.001 | 0.48 (0.39, 0.58) | <.001 | 0.39 (0.23, 0.56) | <.001 |
| >12,500 | 1,332/204 | 0.43 (0.34, 0.51) | <.001 | 0.44 (0.34, 0.54) | <.001 | 0.34 (0.15, 0.52) | <.001 |
| **Fast stepping (minutes/day)** |  |  |  |  |  |  |  |
| ≤5 | 1,245/419 | 1 |  | 1 |  | 1 |  |
| 5-15 | 3,119/660 | 0.17 (0.11, 0.24) | <.001 | 0.12 (0.04, 0.19) | .003 | 0.29 (0.16, 0.41) | <.001 |
| 15-25 | 2,177/390 | 0.27 (0.2, 0.34) | <.001 | 0.24 (0.16, 0.32) | <.001 | 0.32 (0.17, 0.46) | <.001 |
| 25-35 | 1,215/219 | 0.28 (0.2, 0.36) | <.001 | 0.26 (0.17, 0.35) | <.001 | 0.30 (0.13, 0.47) | <.001 |
| >35 | 1,615/289 | 0.25 (0.18, 0.33) | <.001 | 0.23 (0.15, 0.32) | <.001 | 0.26 (0.1, 0.42) | .001 |
| **Peak 1-min cadence (steps/min)** |  |  |  |  |  |  |  |
| ≤100 | 1,222/445 | 1 |  | 1 |  | 1 |  |
| 100-110 | 2,684/647 | 0.18 (0.12, 0.25) | <.001 | 0.19 (0.12, 0.27) | <.001 | 0.14 (0.02, 0.26) | .026 |
| 110-120 | 3,111/598 | 0.30 (0.24, 0.37) | <.001 | 0.31 (0.23, 0.39) | <.001 | 0.27 (0.15, 0.40) | <.001 |
| 120-130 | 1,638/224 | 0.34 (0.27, 0.42) | <.001 | 0.36 (0.27, 0.45) | <.001 | 0.28 (0.11, 0.45) | .001 |
| >130 | 716/63 | 0.38 (0.28, 0.47) | <.001 | 0.40 (0.30, 0.51) | <.001 | 0.21 (-0.06, 0.49) | .129 |
| **Peak 30-min cadence (steps/min)** |  |  |  |  |  |  |  |
| ≤80 | 3,312/933 | 1 |  | 1 |  | 1 |  |
| 80-90 | 2,359/431 | 0.19 (0.14, 0.24) | <.001 | 0.19 (0.13, 0.25) | <.001 | 0.19 (0.07, 0.31) | .001 |
| 90-100 | 1,944/349 | 0.19 (0.13, 0.24) | <.001 | 0.20 (0.14, 0.26) | <.001 | 0.11 (-0.02, 0.24) | .084 |
| 100-110 | 1,062/178 | 0.19 (0.12, 0.26) | <.001 | 0.18 (0.11, 0.26) | <.001 | 0.24 (0.07, 0.4) | .005 |
| >110 | 694/86 | 0.23 (0.15, 0.31) | <.001 | 0.26 (0.17, 0.35) | <.001 | 0.07 (-0.16, 0.3) | .543 |

- 1 Estimates and 95% confidence intervals (CI) for association between step metrics and overall quality of life (z-score). Estimates correspond to Cohen's D effect - 2 sizes. For example, someone from the total population taking 5,000-7,500 steps/day had a 0.30 SD (95% CI: 0.23-0.39) higher overall quality of life compared - 3 to someone taking ≤5,000 steps/day.

### Interaction between step metrics and CVD health status

Associations of steps metrics with overall- and physical QoL were present in individuals with and without CVD (**Figure 3-4**), with similar patterns in both groups for step volume. The dose-response association between time spent fast stepping and QoL was dependent on CVD health status with a significant interaction effect for overall QoL and physical QoL (**Supplementary Table 6**). Slightly lower optimal doses for overall QoL (4 *versus* 9 minutes/day) and physical QoL (6 *versus* 11 minutes/day) were found in individuals with *versus* without CVD, while effect sizes were comparable (CVD: +0.20 SD, 95%CI: 0.12-0.28 *versus* non-CVD; +0.19 SD, 95%CI: 0.12-0.25 for overall QoL, **Figure 4A**; and for physical QoL, CVD: +0.44 SD, 95%CI: 0.28-0.61, *versus* non-CVD: +0.36 SD, 95%CI: 0.28-0.44, **Figure 4D**).

**Figure 3.**
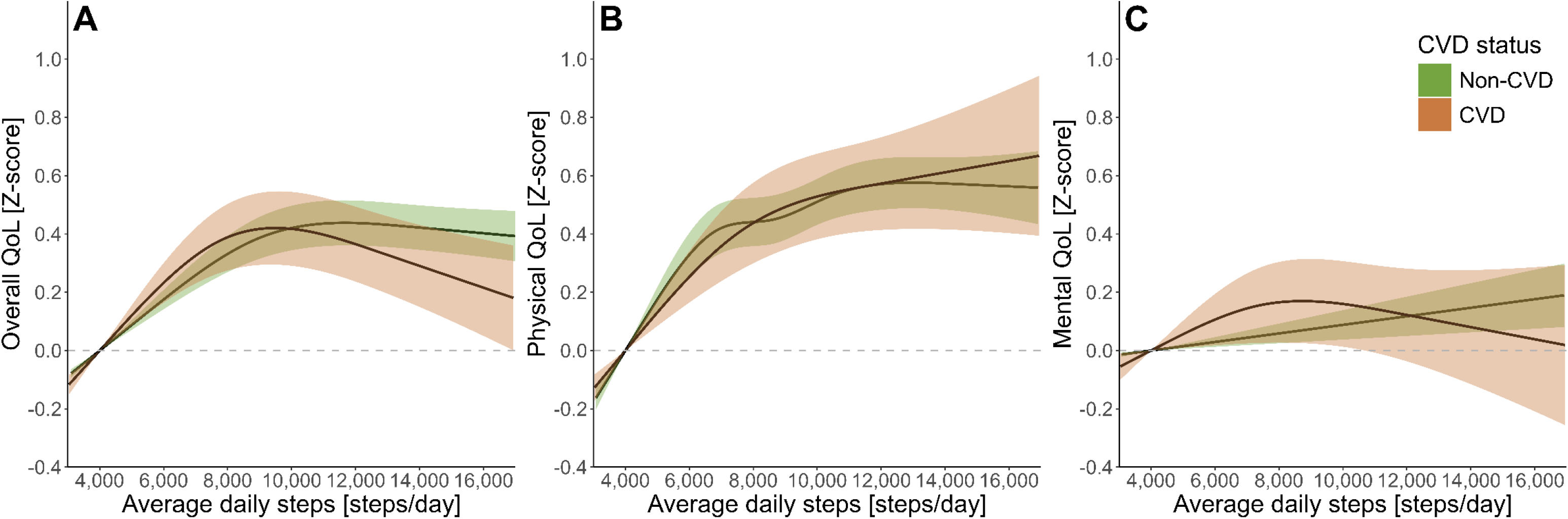
Dose-response association steps and overall, physical and mental quality of life stratified by cardiovascular disease status. Reference values are set at 4,000 steps/day. Shading represents the 95% confidence interval. The model adjusted for age, sex, body mass index, alcohol consumption, smoking status and diabetes. CVD: cardiovascular disease, QoL: quality of life.

**Figure 4.**
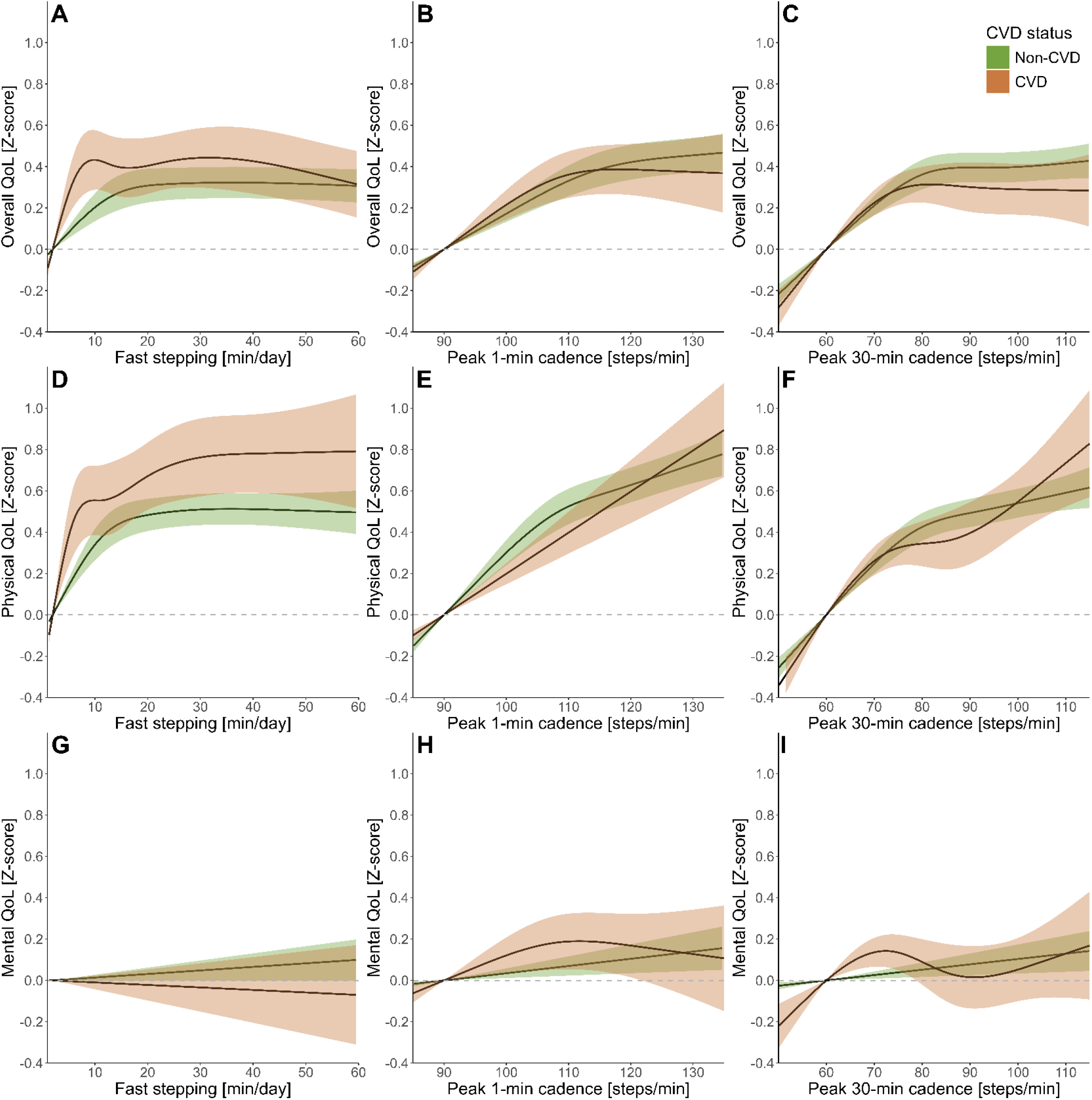
Dose-response association fast stepping (A, D, G), peak 1-min cadence (B, E, H) and peak 30-min cadence (C, F, I) and overall, mental and physical quality of life stratified by cardiovascular disease status. Reference values are set at 2 minutes fast stepping, 90 minutes for peak 1-min cadence, 60 minutes for peak 30-min cadence. Shading represents the 95% confidence interval. The model was adjusted for age, sex, body mass index, alcohol consumption, smoking status and diabetes. CVD: cardiovascular disease, QoL: quality of life.

Similarly, a lower optimum peak 30-min cadence (83 *versus* 93 steps/minute) was found in individuals with *versus* without CVD at a comparable effect size (+0.45 SD, 95% CI: 0.39-0.52 *versus* +0.43 SD, 95% CI: 0.28-0.58) for physical QoL (**Figure 4F**). The other cadence metrics showed similar dose-response curves across groups. The association between step metrics and mental QoL was comparable between individuals with CVD and non-CVD (p for interaction >0.05).

### Impact of QoL instrument and cohort

Sensitivity analyses showed that QoL instrument and cohort did not significantly affect the association between steps/day and QoL (**Supplementary Figure 8**). Other factors that were tested for interaction (age, sex, BMI, CVD type) and removing individuals with other chronic diseases did not affect our inferences (**Supplementary Figure 9-10**).

## Discussion

We showed curvilinear dose-response associations between step metrics and overall- and physical QoL. The curvilinearity means that higher step volume- and intensity metrics are associated with higher QoL, especially at the lower end of the activity spectrum, which subsequently plateau when engaging in large volumes of steps per day. The optimal step volume was observed at 6,561 steps/day, 9 minutes fast stepping/day, 107 steps/minute for peak 1-min cadence and 74 steps/minute for peak 30-min cadence for overall QoL. We found larger effect sizes for the associations of step metrics with physical-compared to mental QoL. Associations for those with and without CVD were similar, except for volume of fast stepping with the optimum at a slightly lower dose in CVD compared to those without CVD (4 *versus* 9 min/day). Taken together, our findings suggest that more and faster steps are associated with better QoL, with optimal QoL benefits at feasible stepping targets.

We found positive and curvilinear dose-response associations between step volume and overall QoL, independent of CVD status. Our findings reinforce observations from general population studies using self-reported PA [35, 36], supporting the concept that more PA is associated with better QoL. Being one of the first studies using accelerometry, we were able to better define and evaluate this curvilinear relation between step metrics and QoL, and to evaluate the interaction with CVD health status. Specifically, we identified an optimal volume of 6,561 steps/day and a maximal effect at 12,427 steps/day for overall QoL. This means that steps beyond 6,561 steps/day are associated with higher QoL, but these volumes are not statistically different compared to the optimum dose. The optimum that we found is slightly lower than the number of daily steps that were previously associated with significant risk reductions for adverse health outcomes (i.e. 7,000 steps/day)[37] and incident CVD (i.e. 7,126 steps/day)[32], but higher than the 5,000 steps in a study into steps and QoL in heart failure patients (n=425).[38] The difference with the latter study may partially be due to a difference in study sample (individuals with heart failure *versus* those with and without CVD) and assessment of steps (smartphone-based *versus* thigh-worn accelerometry). Nevertheless, our findings align well with the recent recommendation of 7,000 steps/day that was proposed by the Australian 24-Hour Movement Guidelines for healthy (older) adults.[39] These step targets are much more feasible than the previously assumed 10,000 steps/day goals for patients with CVD and the population at large.

Stepping cadence matters, as more minutes of fast stepping were associated with a better overall QoL in a curvilinear fashion. We found a significant interaction with CVD health status, with a lower optimal dose in individuals with CVD (4 min/day) compared to individuals without CVD (9 min/day). These observations support the notion that higher-intensity activities yield similar benefits for lower volumes compared to lower-intensity activities.[40] Such low-volume but higher-intensity targets could be achieved by vigorous intensity lifestyle physical activities (VILPA); short bursts of intense physical activity incorporated in daily life like stair climbing, fast walking or short walking sprints up to one minute,[41] or the analogous planned equivalent exercise snacks.[42] Recent studies have shown that exercise snacks can improve cardiorespiratory fitness and lower the risk for adverse health outcomes (i.e. major adverse cardiovascular events, cancer, mortality) in the general population.[42-44] Ultimately, better QoL induced by time spent at fast stepping could improve survival.[45, 46]

Vigorous PA may not be safe or feasible for some patients with CVD, as highlighted in clinical guidelines.[47] It was, therefore, reassuring to observe that optimum QoL values were achieved at a peak 30-min cadence of 74 steps/minute. Such walking pace corresponds to light-intensity PA in healthy individuals[30, 48], but may be perceived as moderate-intensity or higher by people with chronic CVD, depending on their physical fitness level.[49] Our findings highlight that 30 minutes at a relatively low cadence of 74 steps/minute accrued across the day, might already be sufficient to induce significant QoL benefits. Depending on the physical fitness, available time and motivation of the individual, personal step volume and intensity targets could be selected to improve QoL.

We found stronger effects of step metrics on physical-compared to mental QoL. Our observation aligns with previous studies, as stronger effects of PA on physical-*versus* mental QoL are consistently reported across studies in women[35], healthy older adults[50, 51] and individuals with heart failure.[52] Other lifestyle interventions such as mindfulness, or other psychological interventions e.g. embedded in cardiac rehabilitation programs, could be considered as alternatives to improve mental QoL, especially in patients with CVD [53, 54] since psychological intervention effects are potentially larger in clinical populations than in healthy individuals.[55] Future studies need to determine what combination of interventions might improve mental QoL in people with chronic CVD, beyond cardiac rehabilitation.

Strengths of this study include the relatively large study sample (n=9,371) including 1,977 individuals with CVD and the use of device-based step metrics. Our study also had some limitations. First, due to our observational and cross-sectional study design, there is a risk of reverse causation. For example, individuals with CVD may experience a lower QoL than healthy individuals, and because of their lower QoL, may be less active (rather than the other way around). Longitudinal studies are needed to determine the causal relationship between step metrics and QoL. Second, QoL was standardized making the clinical interpretation challenging, as questionnaires may target distinct elements of QoL. However, we deemed this necessary since QoL was assessed with different instruments (i.e., generic versus disease-specific QoL) across studies. Stratified analyses per instrument showed that the associations did not differ between QoL instruments (**Supplementary Figure 8A**). By using standardized QoL, our results allow us to compare with other studies using different QoL instruments. Third, QoL scores were high in both the CVD and non-CVD groups. We therefore expect that the effect sizes we found could be larger in individuals with low-to-moderate QoL. Finally, there is risk of selection bias. Participants in the NES and FACTS cohorts were volunteers from the general population with oversampling of individuals with CVD and The Maastricht Study oversampled individuals with type 2 diabetes. We complemented these cohort data with data from randomized controlled trials in individuals with CVD and consider our dataset as close to the general population as possible and well-suited to answer our research question.

In conclusion, we found that the optimal volume of steps associated with overall QoL is ±6,500 steps/day independent of CVD status. Effect sizes were larger for physical than mental QoL. Both groups demonstrated optimal QoL benefits at feasible stepping targets, with a slightly lower optimum minutes of fast stepping for individuals with CVD.

## Acknowledgements

We sincerely thank all participants in the studies and all the personnel who contributed to the data collection.

## Data availability statement

Metadata (e.g. study protocol, data dictionary) of the STEP COACH project has open access availability via the Open Science Framework with the following digital object identifier (DOI: doi.org/10.17605/OSF.IO/V53AR). Individual participant data of the individual studies will not be made open access for privacy reasons. Individual data will be available under restricted access following the "as open as possible, as closed as necessary" principle. Access to the STEP COACH data can be requested via the PI of the study (dr. Eijsvogels). Following this procedure, this study fully complies with the Open Science principles including FAIR data management, reproducibility, and inclusive, collaborative practices.

