## Supplement 1 for "Accelerometer-derived Step Metrics and Quality of Life in Individuals with and without Cardiovascular Diseases"

Supplemental File 1. Supplementary Tables and Figures

**Supplementary Table 1. Overview of the included studies.**

**Supplementary Table 2. Harmonization of covariates across studies.**

**Supplementary Table 3. Quality of life scores grouped for cohorts using the same instrument stratified by cardiovascular health status.**

**Supplementary Table 4. The associations between categories of step metrics with physical quality of life for the total population and stratified by cardiovascular disease status.**

**Supplementary Table 5. The associations between categories of step metrics with mental quality of life for the total population and stratified by cardiovascular disease status.**

**Supplementary Table 6. P values for interaction effects fast stepping and peak 30-min cadence with cardiovascular health status.**

**Supplementary Figure 1. Example on how to determine the optimal dose.**

**Supplementary Figure 2. Flow-chart of available data.**

**Supplementary Figure 3. Histograms of the frequency of step metrics stratified by cardiovascular disease status.**

**Supplementary Figure 4. Association of quintiles of steps/day and overall, physical and mental quality of life for the overall population.**

**Supplementary Figure 5. Association of quintiles of fast stepping, peak 1-min cadence and peak 30-min cadence and overall, physical and mental quality of life for the overall population.**

**Supplementary Figure 6. Association of quintiles of steps and overall, physical and mental quality of life stratified by cardiovascular disease status.**

**Supplementary Figure 7. Association of quintiles of fast stepping, peak 1-min cadence and peak 30-min cadence and overall, physical and mental quality of life stratified by cardiovascular disease status.**

**Supplementary Figure 8. Dose-response association steps and overall quality of life.**

**Supplementary Figure 9. Dose-response association steps and overall quality of life.**

**Supplementary Figure 10. Dose-response association steps and overall quality of life.**

**Supplementary Table 1. Overview of the included studies.**

| Cohort | Sample decription, age and sample size | Study type | Timing assessment vs CV event | CVD ascertain-ment | Quality of life instrument | Leading institutution, country | Eligibility criteria | Ethics approval (committee name, reference number) |
|---|---|---|---|---|---|---|---|---|
| The Physical Activity in Statin Users Study [1] | Adults from different general practices in the Netherlands, currently or previously on statins, ≥ 18 years, n=194 | Longitudinal cohort | Chronic CVD | Self-report | 1 item (1-10) | Radboud university medical center, Netherlands | Individuals ≥ 18 years, on statin therapy or on statin therapy in the 5 years prior to participation, no other diseases known to cause muscle symptoms | The Local Ethics Committee on Research Involving Human Subjects (CMO) of the region Arnhem and Nijmegen, the Netherlands (NL9196) |
| Nijmegen Exercise Study [2] | Participants in Nijmegen Four days Marches or Seven Hills Run, and their friends & family members, ≥ 18 years, n=2,007 | Longitudinal cohort | Chronic CVD | Self-report | 1 item (1-10) | Radboud university medical center, Netherlands | Individuals of the general population participating in Dutch sport events (i.e. International Nijmegen Four Days Marches and the Seven Hills Run) and their family and friends. Pregnant individuals are excluded. | The Local Ethics Committee on Research Involving Human Subjects (CMO) of the region Arnhem and Nijmegen, the Netherlands (NL36743.091.11) |
| The Maastricht Study [3] | General population with an oversampling of those with type 2 diabetes, 40-75 years, n=7,704 | Longitudinal cohort | Chronic CVD | Self-report | SF-36v1 | Maastricht University Medical Center+ and Maastricht University, Netherlands | Individuals aged 40-75 years, oversampling of those with type 2 diabetes | Institutional medical ethical committee (NL31329.068.10) and the Minister of Health, Welfare and Sports of the Netherlands (Permit 131088-105234-PG) |

| | | | | | | | | |
|---|---|---|---|---|---|---|---|---|
| Cardiac RehApp [4] | Patients with CAD referred to a CR program, >18 years, n=37 | RCT including two intervention arms: mHealth + CR vs usual CR | 12 wks post event | Electronic patient file | HeartQoL | Radboud university medical center, Netherlands | Individuals >18 years, diagnosed with coronary artery disease, able to use a smartphone, in the possession of a smartphone with internet access at home, referred to cardiac rehabilitation, able to understand and perform study procedures, no contraindications to exercise, no severe orthopaedic problems restricting physical activity, and no significant language barrier | The Local Ethics Committee on Research Involving Human Subjects (CMO) of the region Arnhem and Nijmegen, the Netherlands (NL72182.091.019) |
| SIT LESS [5] | Patients referred to CR because of (in)stable angina, MI and/or after coronary revascularization, ≥ 18 years, n=178 | RCT with two intervention arms: BCT + CR vs usual CR | 6 mo post event | Electronic patient file | HeartQoL | Radboud university medical center, Netherlands | Individuals ≥18 years, undergoing CR because of (in)stable angina, MI and/or coronary revascularization, be able to understand and perform study procedures, NYHA class <3, able to stand/walk, no expected CABG within 8 weeks of inclusion, | The Local Ethics Committee on Research Involving Human Subjects (CMO) of the region Arnhem and Nijmegen, the Netherlands (NL72604.091.20) |

| | | | | | | | |
|---|---|---|---|---|---|---|---|
| | | | | | | | no participation in another trial on physical activity at the same time |

BCT: behavior change techniques, CABG: coronary artery bypass grafting, CAD: coronary artery disease, CR: cardiac rehabilitation, CVD: cardiovascular disease, CV: cardiovascular, HC: healthy controls, MI: myocardial infarct, NYHA: New York Heart Association, RCT: randomized controlled trial, SF-36v1: Short-Form 36 version 1.

**Supplementary Table 2. Harmonization of covariates across studies.**

| Covariate | Harmonized construct | SIT LESS | Cardiac RehApp | The Maastricht Study | The Physical Activity in Statin Users Study | Nijmegen Exercise Study |
|---|---|---|---|---|---|---|
| Age | Continuous (years) | Age at admission (years) | Age at admission (years) | Age at visit (years) | Difference between date of birth and inclusion date | Difference between date of birth and inclusion date, if NA based on questionnaire |
| Sex | 1: Female<br>2: Male | 1: Female<br>2: Male | 1: Female<br>2: Male | 1: Female<br>2: Male | 1: Female<br>2: Male | 1: Female<br>2: Male |
| BMI | Continuous (kg/m$^2$) | Questionnaire at inclusion<br>Height: cm<br>Weight: kg | Questionnaire at inclusion<br>Height: m<br>Weight: kg | Objectively measured<br>Height: cm<br>Weight: kg | Questionnaire at inclusion<br>Height: cm<br>Weight: kg | Objectively measured<br>Height: cm<br>Weight: kg |
| Alcohol use | Current alcohol use<br>0: No<br>1: Yes | **Question:** "Do you drink alcohol?"<br><br>**Coding:**<br>0: No, never; No, stopped<br>1: Yes | **Question:** "Do you drink alcohol?"<br><br>**Coding:**<br>0: No<br>1: Yes | **Question:** "Do you drink alcohol?"<br><br>**Coding:**<br>0: No<br>1: Yes | **Question:** "How many glasses of beer/wine do you drink on average per week?"<br><br>**Coding:**<br>0: 0 glasses/week<br>1: ≥1 glass/week | **Question:** "What is your current alcohol consumption status?"<br><br>**Coding:**<br>0: Never; Former<br>1: Current |
| Smoking status | Current smoking<br>0: No<br>1: Yes | **Question**: "Do you smoke?"<br><br>**Coding:**<br>0: No, never; No, stopped<br>1: Yes | **Question:** "Do you smoke?"<br><br>**Coding:**<br>0: No<br>1: Yes | **Question:** "Do you smoke?"<br><br>**Coding:**<br>0: Never; Former<br>1: Current | **Question:** "Do you smoke?"<br><br>**Coding:**<br>0: No, never; No, former<br>1: Yes, current | **Question:** "Do you smoke?"<br><br>**Coding:**<br>0: No, never; No, former<br>2: Yes, current |
| Diabetes mellitus | 0: No<br>1: Yes | Obtained from electronic patient file<br><br>**Coding:** | Obtained from electronic patient file. | Type 2 diabetes status according to oral glucose tolerance test and medication use | **Question:** "Did you regularly use anti diabetic | **Question:** "Which of the following diseases below has been diagnosed by a physician? Diabetes" |

| | | | | | | |
|---|---|---|---|---|---|---|
| | | 0: No<br>1: Yes | **Coding:**<br>0: No<br>1: Yes | incl. other types of diabetes<br><br>**Coding:**<br>0: No diabetes; Prediabetes<br>1: Type 2 diabetes; Other type of diabetes | medication/insulin in the past year?"<br><br>**Coding:**<br>0: No<br>1: Yes | **Coding:**<br>0: No<br>1: Yes |
| **Cardiovascular health status** | | Obtained from electronic patient file | Obtained from electronic patient file | Obtained from questionnaires | Obtained from questionnaires | Obtained from questionnaires |
| Myocardial Infarct | 0: No<br>1: Yes | **Coding:**<br>0: No<br>1: Yes | **Coding:**<br>0: No<br>1: Yes | **Rose Questionnaire Coding:**<br>0: No<br>1: Yes | **Question:** "Which type of diseases below has been diagnosed by physician? Myocardial infarction"<br><br>**Coding:**<br>0: No<br>1: Yes | **Question:** "Which type of diseases below has been diagnosed by physician? Myocardial infarction"<br><br>**Coding:**<br>0: No<br>1: Yes |
| Angina pectoris with use of medication | 0: No<br>1: Yes | **Coding:**<br>0: No<br>1: Yes | **Coding:**<br>0: No<br>1: Yes | **Rose Questionnaire Coding:**<br>0: No; Probably<br>1: Yes | NA | NA |
| Percutaneous coronary intervention or other surgery on heart vessels | 0: No<br>1: Yes | **Coding:**<br>0: No<br>1: Yes | **Coding:**<br>0: No<br>1: Yes | **Question:** "Which vessels did you have surgery on? Heart"; "Which vessels were opened with angioplasty? Heart vessels"<br>0: No<br>1: Yes | NA | NA |

| | | | | | | |
|---|---|---|---|---|---|---|
| Heart failure | 0: No<br>1: Yes | **Coding:**<br>0: No<br>1: Yes | **Coding:**<br>0: No<br>1: Yes | NA | **Question:** "Which type of diseases below has been diagnosed by physician? Heart failure"<br><br>**Coding:**<br>0: No<br>1: Yes | **Question:** "Which type of diseases below has been diagnosed by physician? Heart failure"<br><br>**Coding:**<br>0: No<br>1: Yes |
| Rhythm disorder | 0: No<br>1: Yes | **Atrial fibrillation**<br>0: No<br>1: Yes | **Atrial fibrillation**<br>0: No<br>1: Yes | NA | NA | **Question:** "Which type of diseases below has been diagnosed by physician? Atrial fibrillation"<br><br>0: No<br>1: Yes |
| Heart valve diseases | 0: No<br>1: Yes | **Coding:**<br>0: No<br>1: Yes | **Coding:**<br>0: No<br>1: Yes | NA | NA | NA |
| Peripheral artery disease | 0: No<br>1: Yes | **Coding:**<br>0: No<br>1: Yes | **Coding:**<br>0: No<br>1: Yes | NA | **Question:** "Which type of diseases below has been diagnosed by physician? Thrombosis"<br><br>0: No<br>1: Yes | **Question:** "Which type of diseases below has been diagnosed by physician? Thrombosis"<br><br>0: No<br>1: Yes |
| Cerebrovascular accident | 0: No<br>1: Yes | **Coding:**<br>0: No; TIA<br>1: Yes | **Coding:**<br>0: No<br>1: Yes | **Rose Questionnaire Coding:**<br>0: No<br>1: Yes | **Question:** "Which type of diseases below has been diagnosed by physician? Stroke"<br><br>**Coding:**<br>0: No | **Question:** "Which type of diseases below has been diagnosed by physician? Stroke"<br><br>**Coding:**<br>0: No |

| | | | | | | |
|---|---|---|---|---|---|---|
| | | | | | 1: Yes | 1: Yes |
| CVD, type unknown | 0: No<br>1: Yes | NA | **Coding:**<br>0: No<br>1: Yes | **Coding:**<br>0: No<br>1: Yes, self-reported CVD but type unspecified | NA | NA |

CVD: cardiovascular disease.

**Supplementary Table 3. Quality of life scores grouped for cohorts using the same instrument stratified by cardiovascular health status.**

| | Cardiac RehApp/SIT LESS | The Maastricht Study | | Nijmegen Exercise Study/ The Physical Activity in Statin Users Study | |
|---|---|---|---|---|---|
| | CVD (n=186) | Non-CVD (n=6,003) | CVD (n=1,285) | Non-CVD (n=1,391) | CVD (n=506) |
| **Overall QoL** | 2.0 [1.6–2.8] | 0.8 [0.8–0.9] | 0.8 [0.7–0.9] | 8.0 [8.0–9.0] | 8.0 [7.0–9.0] |
| **Physical QoL** | 2.1 [1.4–2.8] | 50.9 [44.6–54.7] | 46.1 [36.1–52.2] | NA | NA |
| **Mental QoL** | 2.3 [1.8–3.0] | 55.8 [51.1–59.1] | 55.7 [49.6–59.4] | NA | NA |

Median and interquartile ranges [Q1-Q3] are reported. CVD: cardiovascular disease, QoL: quality of life.

**Supplementary Table 4. The associations between categories of step metrics with physical quality of life for the total population and stratified by cardiovascular disease status.**

| | | Total population | | Non-CVD | | CVD | |
|---|---|---|---|---|---|---|---|
| **Step metric** | **n/CVD** | **Estimate (95% CI)** | **p-value** | **Estimate (95% CI)** | **p-value** | **Estimate (95% CI)** | **p-value** |
| **Steps/day** | | | | | | | |
| ≤5,000 | 782/285 | 1 | | 1 | | 1 | |
| 5,000-7,500 | 2,109/470 | 0.36 (0.28, 0.43) | <.001 | 0.33 (0.24, 0.42) | <.001 | 0.39 (0.23, 0.55) | <.001 |
| 7,500-10,000 | 2,362/412 | 0.46 (0.38, 0.53) | <.001 | 0.44 (0.35, 0.52) | <.001 | 0.47 (0.3, 0.63) | <.001 |
| 10,000-12,500 | 1,464/206 | 0.56 (0.47, 0.64) | <.001 | 0.54 (0.44, 0.63) | <.001 | 0.57 (0.37, 0.78) | <.001 |
| >12,500 | 750/94 | 0.58 (0.48, 0.67) | <.001 | 0.54 (0.44, 0.65) | <.001 | 0.72 (0.46, 0.98) | <.001 |
| **Fast stepping (minutes/day)** | | | | | | | |
| ≤5 | 1,114/348 | 1 | | 1 | | 1 | |
| 5-15 | 2,803/556 | 0.28 (0.21, 0.34) | <.001 | 0.24 (0.17, 0.31) | <.001 | 0.34 (0.19, 0.49) | <.001 |
| 15-25 | 1,850/303 | 0.43 (0.36, 0.5) | <.001 | 0.38 (0.31, 0.46) | <.001 | 0.54 (0.37, 0.71) | <.001 |
| 25-35 | 893/148 | 0.47 (0.38, 0.55) | <.001 | 0.44 (0.35, 0.53) | <.001 | 0.52 (0.31, 0.74) | <.001 |
| >35 | 807/112 | 0.48 (0.39, 0.56) | <.001 | 0.43 (0.34, 0.52) | <.001 | 0.68 (0.45, 0.91) | <.001 |
| **Peak 1-min cadence (steps/min)** | | | | | | | |
| ≤100 | 1,016/336 | 1 | | 1 | | 1 | |
| 100-110 | 2,290/502 | 0.25 (0.19, 0.32) | <.001 | 0.25 (0.18, 0.33) | <.001 | 0.23 (0.08, 0.38) | .003 |
| 110-120 | 2,583/450 | 0.42 (0.35, 0.49) | <.001 | 0.43 (0.35, 0.5) | <.001 | 0.36 (0.21, 0.52) | <.001 |

| | | | | | | | |
|---|---|---|---|---|---|---|---|
| 120-130 | 1,196/148 | 0.51 (0.43, 0.59) | <.001 | 0.51 (0.43, 0.6) | <.001 | 0.46 (0.25, 0.68) | <.001 |
| >130 | 382/31 | 0.64 (0.53, 0.76) | <.001 | 0.63 (0.52, 0.75) | <.001 | 0.84 (0.44, 1.24) | <.001 |
| **Peak 30-min cadence (steps/min)** | | | | | | | |
| ≤80 | 2,929/762 | 1 | | 1 | | 1 | |
| 80-90 | 2,023/326 | 0.23 (0.18, 0.28) | <.001 | 0.22 (0.16, 0.28) | <.001 | 0.26 (0.12, 0.4) | <.001 |
| 90-100 | 1,537/247 | 0.28 (0.22, 0.34) | <.001 | 0.29 (0.22, 0.35) | <.001 | 0.22 (0.06, 0.38) | .006 |
| 100-110 | 699/103 | 0.32 (0.24, 0.4) | <.001 | 0.31 (0.23, 0.39) | <.001 | 0.41 (0.19, 0.63) | <.001 |
| >110 | 279/29 | 0.47 (0.36, 0.59) | <.001 | 0.41 (0.3, 0.53) | <.001 | 0.99 (0.59, 1.39) | <.001 |

Estimates and 95% confidence intervals (CI) for association between step metrics and physical quality of life (z-score). Estimates correspond to Cohen's D effect sizes.

**Supplementary Table 5. The associations between categories of step metrics with mental quality of life for the total population and stratified by cardiovascular disease status.**

| | | Total population | | Non-CVD | | CVD | |
|---|---|---|---|---|---|---|---|
| **Step metric** | **n/CVD** | **Estimate (95% CI)** | **p-value** | **Estimate (95% CI)** | **p-value** | **Estimate (95% CI)** | **p-value** |
| **Steps/day** | | | | | | | |
| ≤5,000 | 782/285 | 1 | | 1 | | 1 | |
| 5,000-7,500 | 2,109/470 | 0.08 (0, 0.17) | .044 | 0.03 (-0.06, 0.13) | .497 | 0.2 (0.04, 0.36) | .016 |
| 7,500-10,000 | 2,362/412 | 0.14 (0.05, 0.22) | .001 | 0.11 (0.02, 0.21) | .022 | 0.15 (-0.02, 0.32) | .082 |
| 10,000-12,500 | 1,464/206 | 0.14 (0.05, 0.23) | .003 | 0.12 (0.01, 0.22) | .027 | 0.12 (-0.08, 0.33) | .227 |
| >12,500 | 750/94 | 0.16 (0.05, 0.26) | .003 | 0.14 (0.03, 0.26) | .015 | 0.12 (-0.13, 0.38) | .350 |
| **Fast stepping (minutes/day)** | | | | | | | |
| ≤5 | 1,114/348 | 1 | | 1 | | 1 | |
| 5-15 | 2,803/556 | 0.03 (-0.04, 0.1) | .409 | -0.01 (-0.09, 0.07) | .861 | 0.12 (-0.02, 0.27) | .099 |
| 15-25 | 1,850/303 | 0.06 (-0.02, 0.13) | .146 | 0.03 (-0.05, 0.12) | .459 | 0.11 (-0.06, 0.28) | .205 |
| 25-35 | 893/148 | 0.07 (-0.02, 0.16) | .112 | 0.06 (-0.04, 0.16) | .219 | 0.07 (-0.14, 0.28) | .531 |
| >35 | 807/112 | 0.05 (-0.04, 0.14) | .310 | 0.04 (-0.06, 0.14) | .429 | 0.03 (-0.21, 0.26) | .833 |
| **Peak 1-min cadence (steps/min)** | | | | | | | |
| ≤100 | 1,016/336 | 1 | | 1 | | 1 | |
| 100-110 | 2,290/502 | 0.06 (-0.01, 0.13) | .115 | 0.05 (-0.04, 0.13) | .299 | 0.08 (-0.07, 0.22) | .317 |

| | | | | | | | |
|---|---|---|---|---|---|---|---|
| 110-120 | 2,583/450 | 0.07 (0, 0.15) | .061 | 0.05 (-0.04, 0.14) | .248 | 0.12 (-0.04, 0.28) | .137 |
| 120-130 | 1,196/148 | 0.13 (0.05, 0.22) | .003 | 0.12 (0.03, 0.22) | .014 | 0.15 (-0.07, 0.37) | .175 |
| >130 | 382/31 | 0.08 (-0.04, 0.2) | .172 | 0.1 (-0.03, 0.22) | .147 | -0.11 (-0.51, 0.29) | .588 |
| **Peak 30-min cadence (steps/min)** | | | | | | | |
| ≤80 | 2,929/762 | 1 | | 1 | | 1 | |
| 80-90 | 2,023/326 | 0.04 (-0.01, 0.1) | .145 | 0.05 (-0.01, 0.12) | .095 | 0 (-0.14, 0.14) | .989 |
| 90-100 | 1,537/247 | 0.06 (0, 0.13) | .044 | 0.08 (0.02, 0.15) | .015 | -0.03 (-0.19, 0.13) | .694 |
| 100-110 | 699/103 | 0.01 (-0.07, 0.09) | .784 | 0.01 (-0.08, 0.1) | .856 | 0.07 (-0.16, 0.29) | .553 |
| >110 | 279/29 | 0.17 (0.05, 0.3) | .005 | 0.2 (0.07, 0.33) | .003 | 0.06 (-0.34, 0.46) | .775 |

Estimates and 95% confidence intervals (CI) for association between step metrics and mental quality of life (z-score). Estimates correspond to Cohen's D effect sizes.

**Supplementary Table 6. P values for interaction effects fast stepping and peak 30-min cadence with cardiovascular health status.**

| | **Fast stepping** | | **Peak 30-min cadence** |
|---|---|---|---|
| | **Overall QoL** | **Physical QoL** | **Physical QoL** |
| Spline 1 | .01 | .02 | .17 |
| Spline 2 | .04 | .06 | .07 |
| Spline 3 | .05 | .08 | .03 |
| Spline 4 | NA | .12 | NA |

P-values for the interaction of step metrics with cardiovascular disease status and overall- and physical QoL. For fast stepping (overall QoL) and peak 30-min cadence (physical QoL) 3 splines are presented for models with 4 knots. For fast stepping (physical QoL) 4 splines are presented for a model with 5 knots. NA: not applicable, QoL: quality of life.

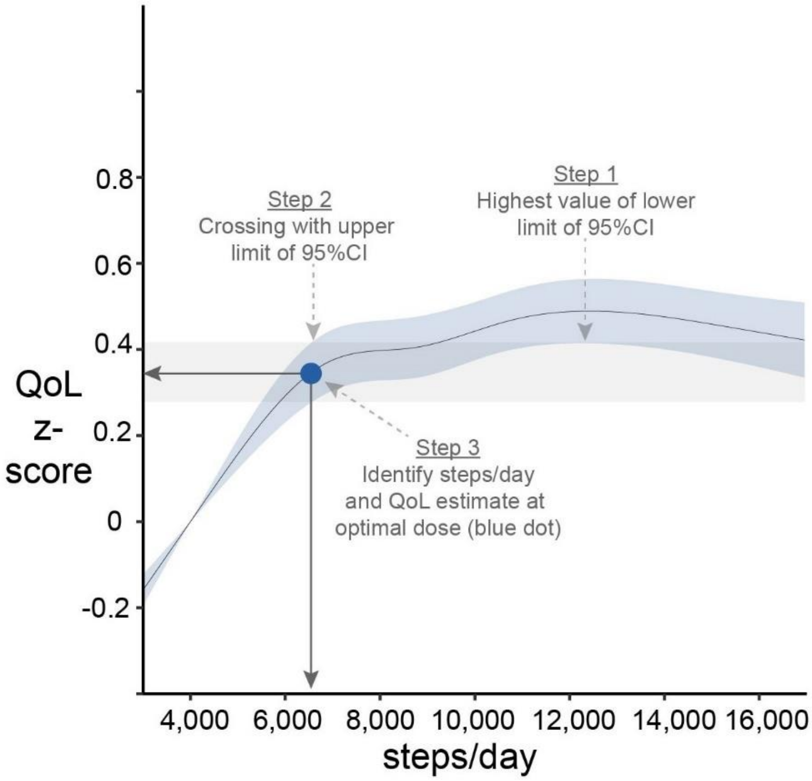

**Supplementary Figure 1. Example on how to determine the optimal dose.** First, identify the highest value of the lower limit of the 95% confidence interval. Second, determine where the lower limit meets the upper limit. Third, identify the steps/day and QoL estimate corresponding to the optimal dose (blue dot). Increasing steps/day beyond this optimum (grey bar) does not yield statistically significant benefits.

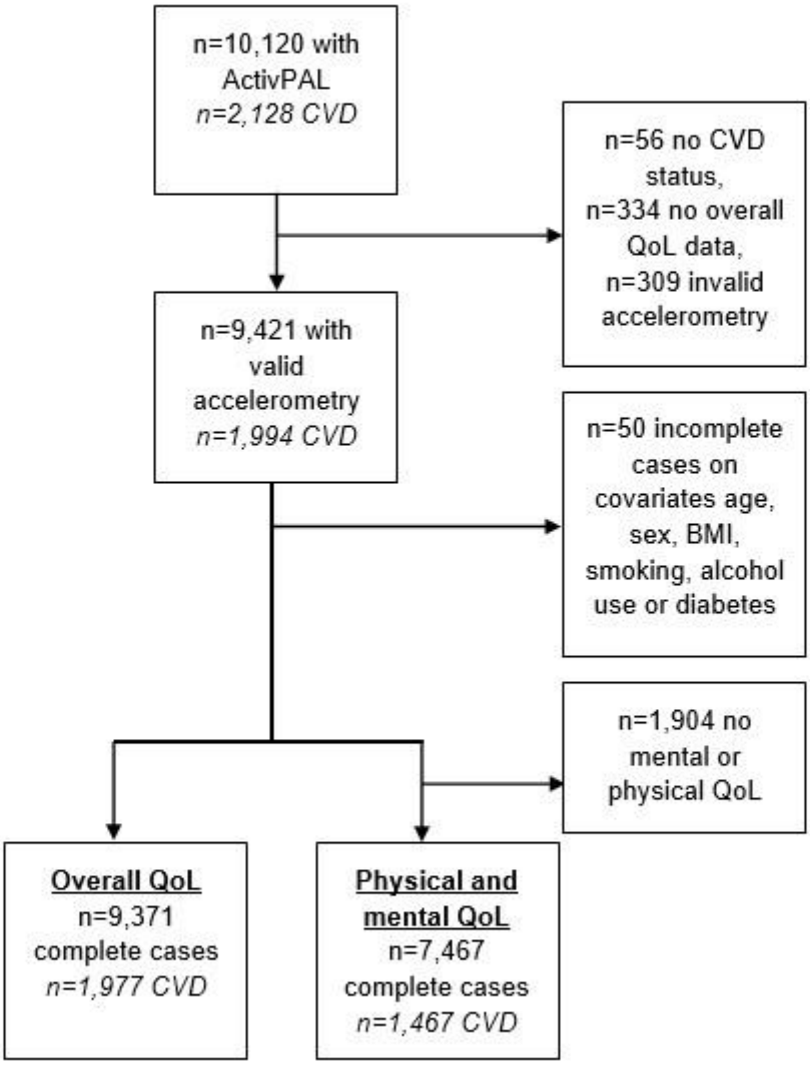

**Supplementary Figure 2. Flow-chart of available data.** CVD: cardiovascular disease, QoL: quality of life.

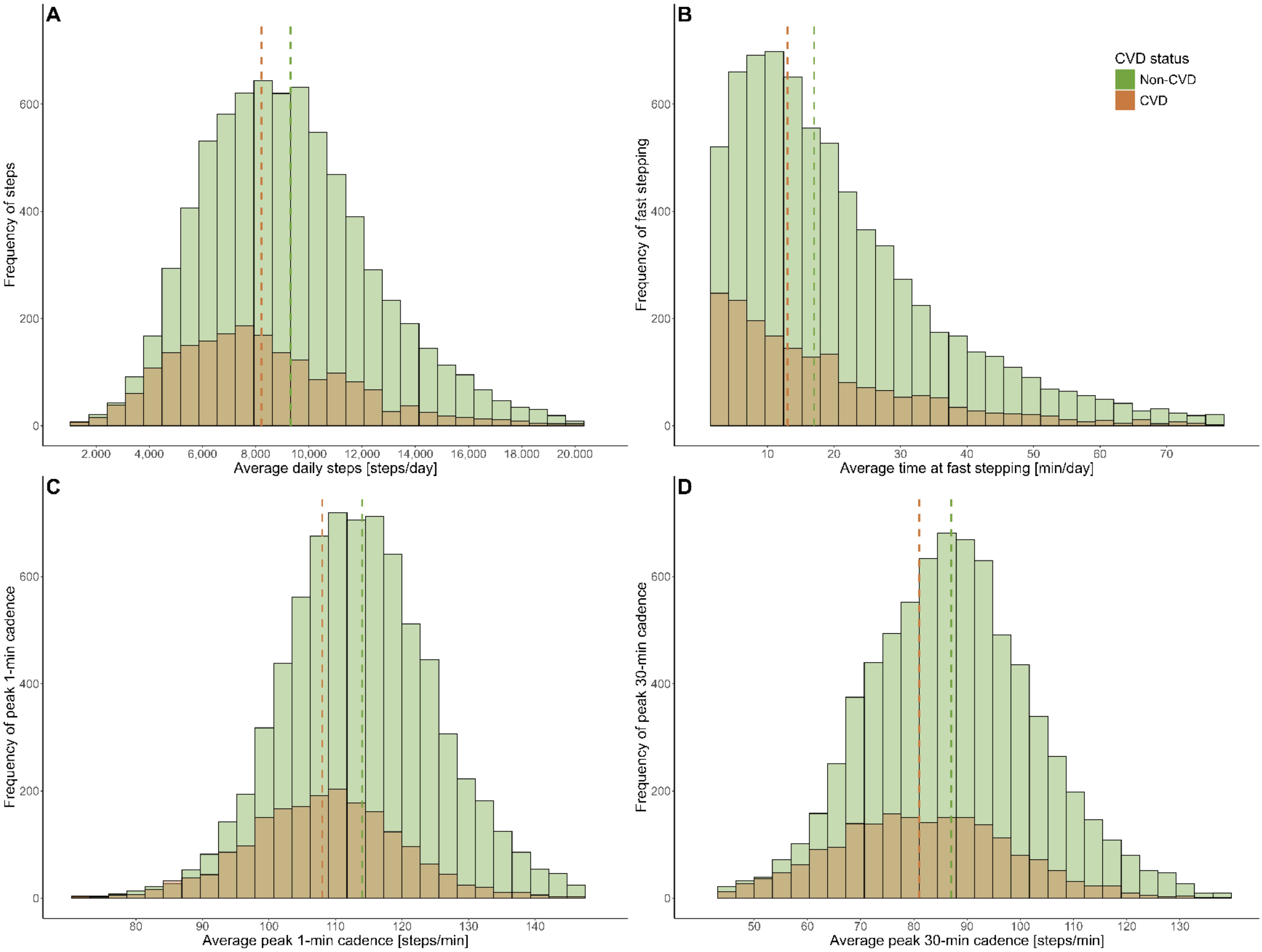

**Supplementary Figure 3. Histograms of the frequency of step metrics stratified by cardiovascular disease status.** Dashed line represents mean step metric per group (median for fast stepping). CVD: cardiovascular disease.

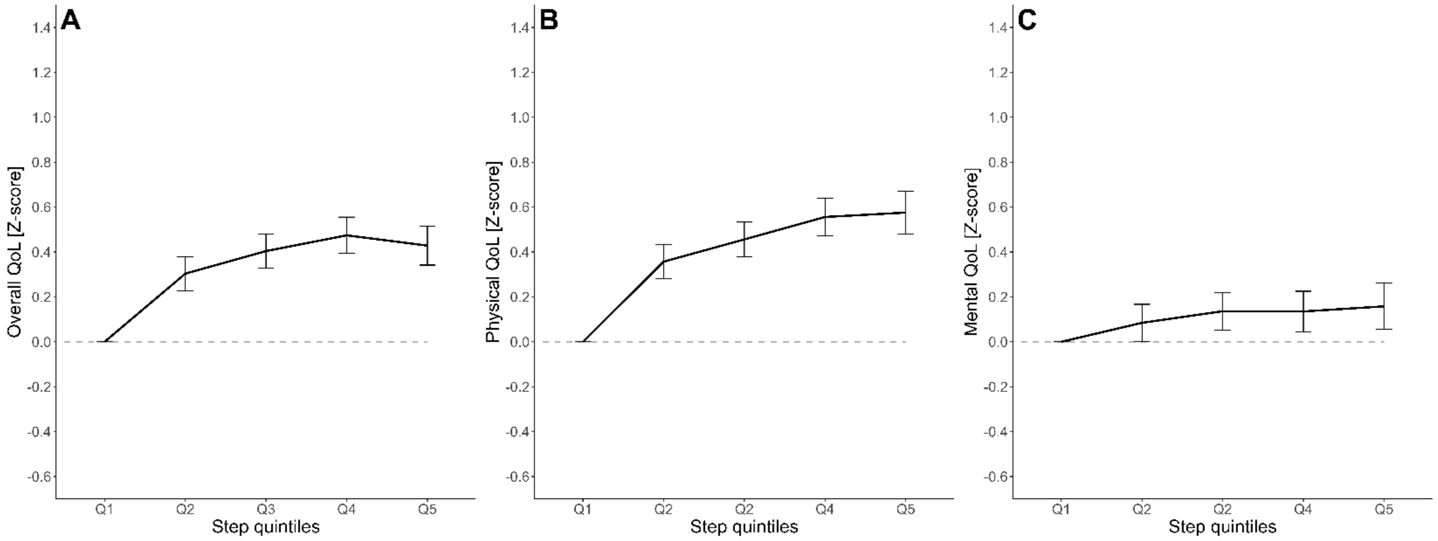

**Supplementary Figure 4. Association of quintiles of steps/day and overall (A), physical (B) and mental (C) quality of life for the overall population.** Q1 is the reference category (≤5,000 steps/day). Bars represent the 95% confidence interval. QoL: quality of life.

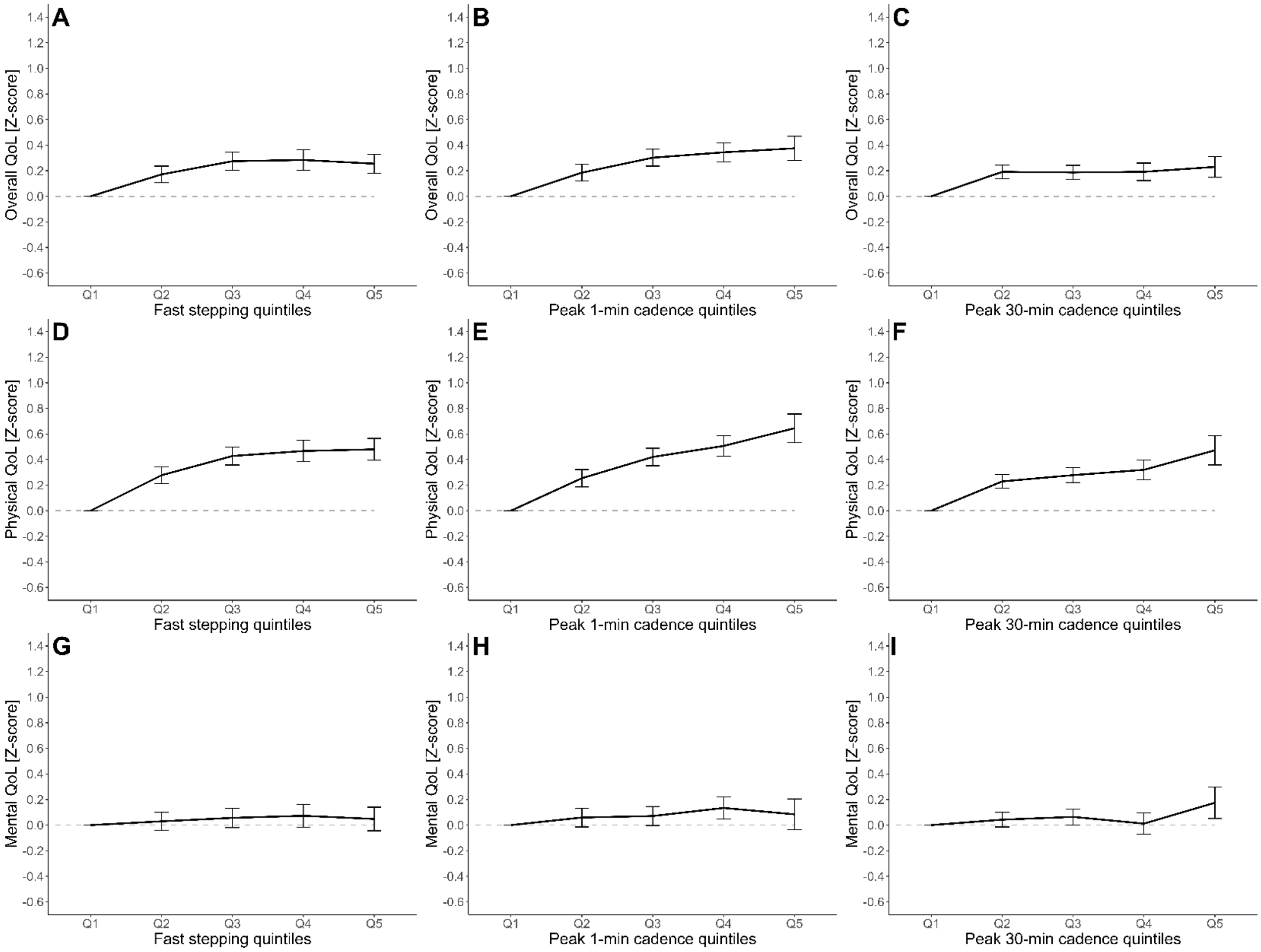

**Supplementary Figure 5. Association of quintiles of fast stepping (A,D,G), peak 1-min cadence (B,E,H) and peak 30-min cadence (C,F,I) and overall, physical and mental quality of life for the overall population.** Q1 is the reference category (≤5,000 steps/day). Bars represent the 95% confidence interval. QoL: quality of life.

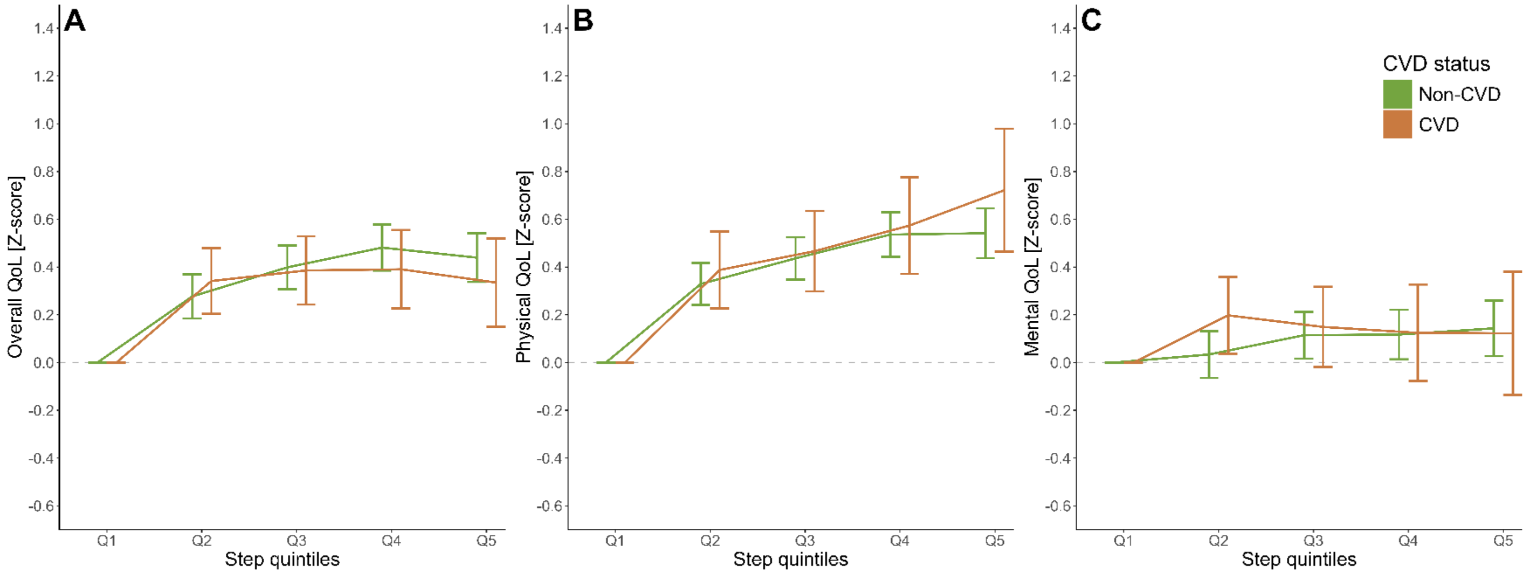

**Supplementary Figure 6. Association of quintiles of steps and overall (A), physical (B) and mental (C) quality of life stratified by cardiovascular disease status.** Q1 is the reference category (≤5,000 steps/day). Bars represent the 95% confidence interval. CVD: cardiovascular disease, QoL: quality of life.

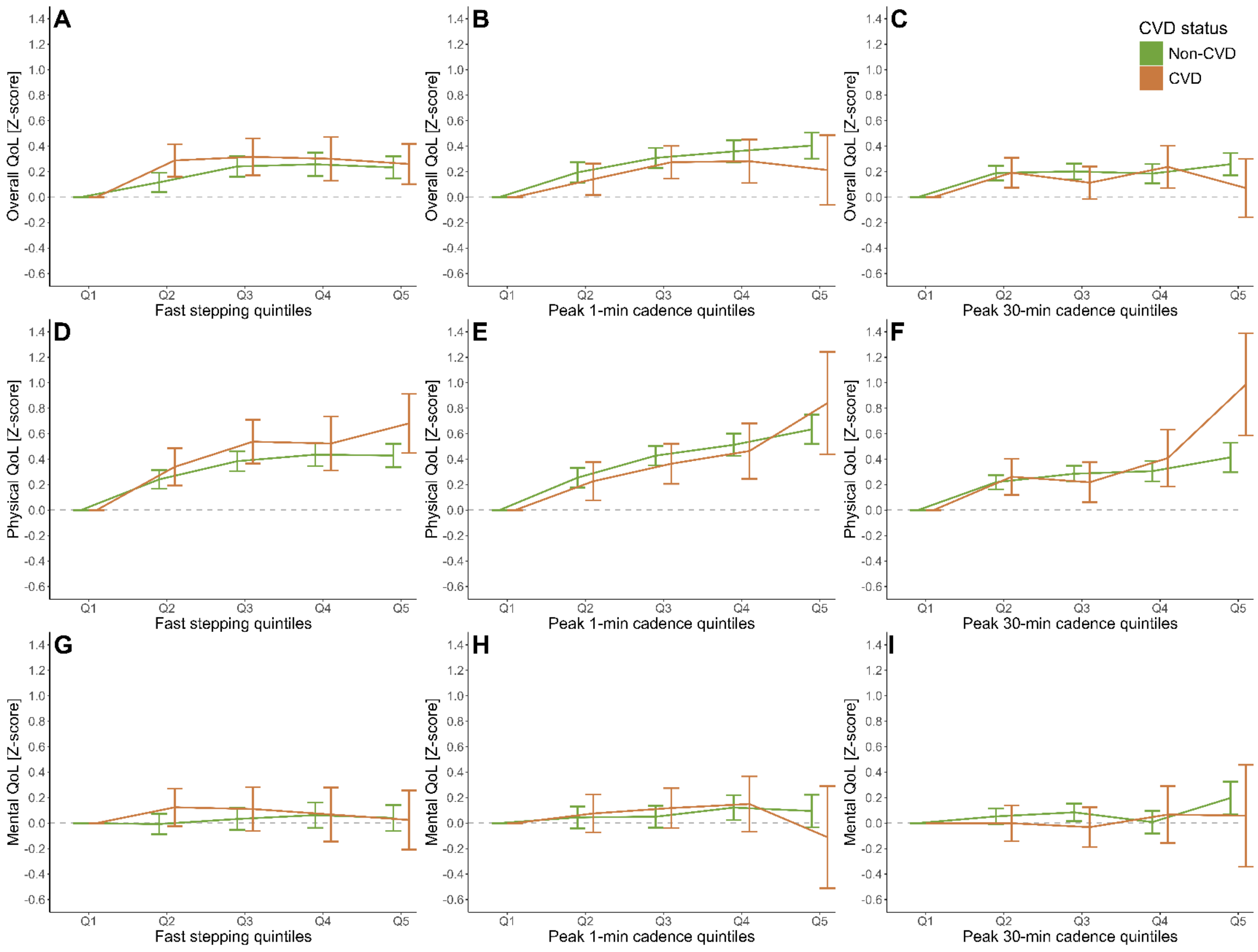

**Supplementary Figure 7. Association of quintiles of fast stepping (A,D,G), peak 1-min cadence (B,E,H) and peak 30-min cadence (C,F,I) and overall, physical and mental quality of life stratified by cardiovascular disease status.** Q1 is the reference category (≤5,000 steps/day). Bars represent the 95% confidence interval. CVD: cardiovascular disease, QoL: quality of life.

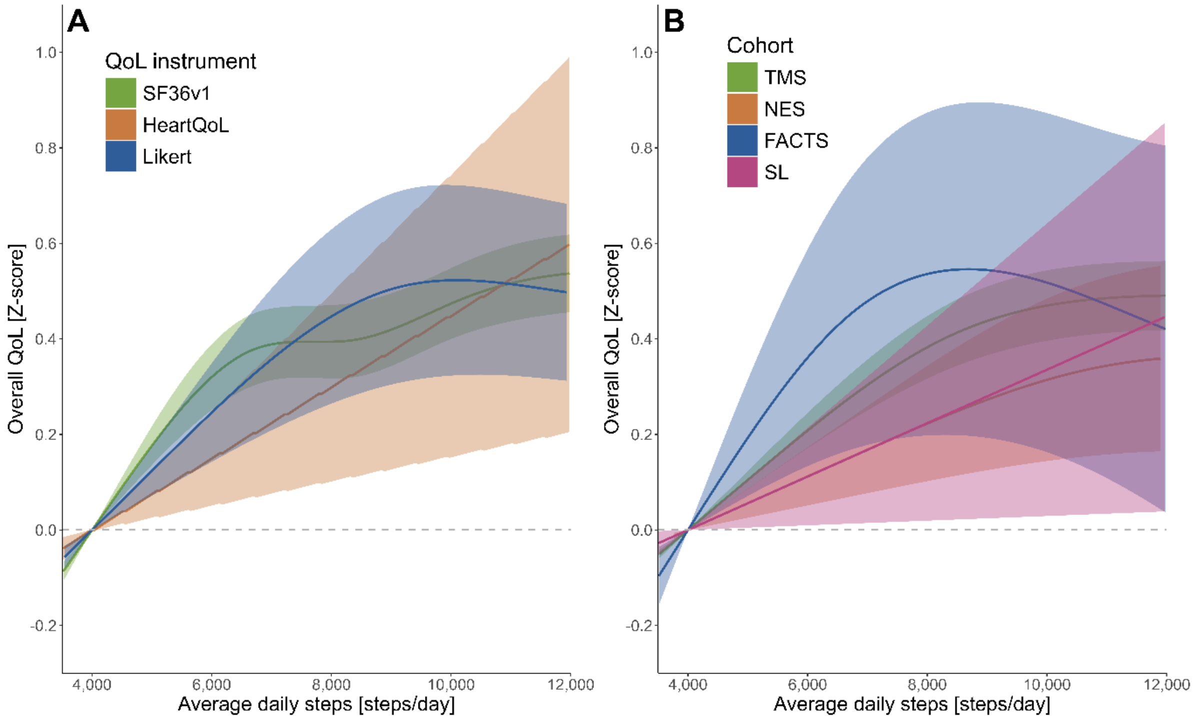

**Supplementary Figure 8. Dose-response association steps and overall quality of life.** The reference value is set at 4,000 steps/day. (A): per QoL instrument. (B): per cohort. Cardiac RehApp was not included in this analysis, since the sample size (n=29) was too small. Shading represents the 95% confidence interval. FACTS: The Physical Activity in Statin Users Study, NES: Nijmegen Exercise Study, SF36v1: Short-Form-36v1, SL: SITLESS, TMS: The Maastricht Study.

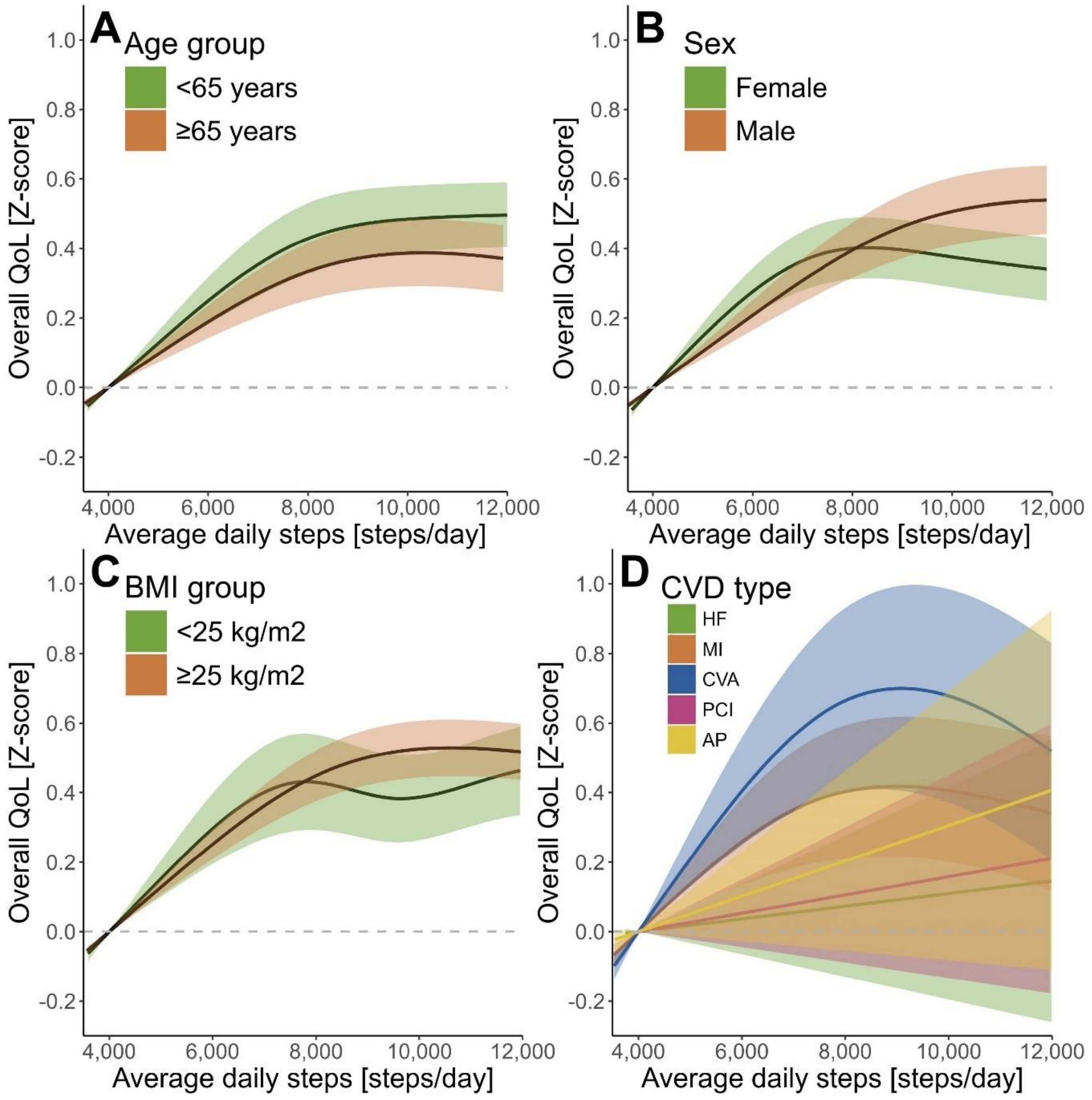

**Supplementary Figure 9**. **Dose-response association steps and overall quality of life.** The reference value is set at 4,000 steps/day. (A): age group <65 and ≥65 years. (B): sex. (C): body mass index <25 kg/m$^2$ versus ≥ 25 kg/m$^2$. (D): per cardiovascular disease type. Shading represents the 95% confidence interval. AP: angina pectoris, CVD: cardiovascular disease, HF: heart failure, MI: myocardial infarct, PCI: percutaneous coronary intervention.

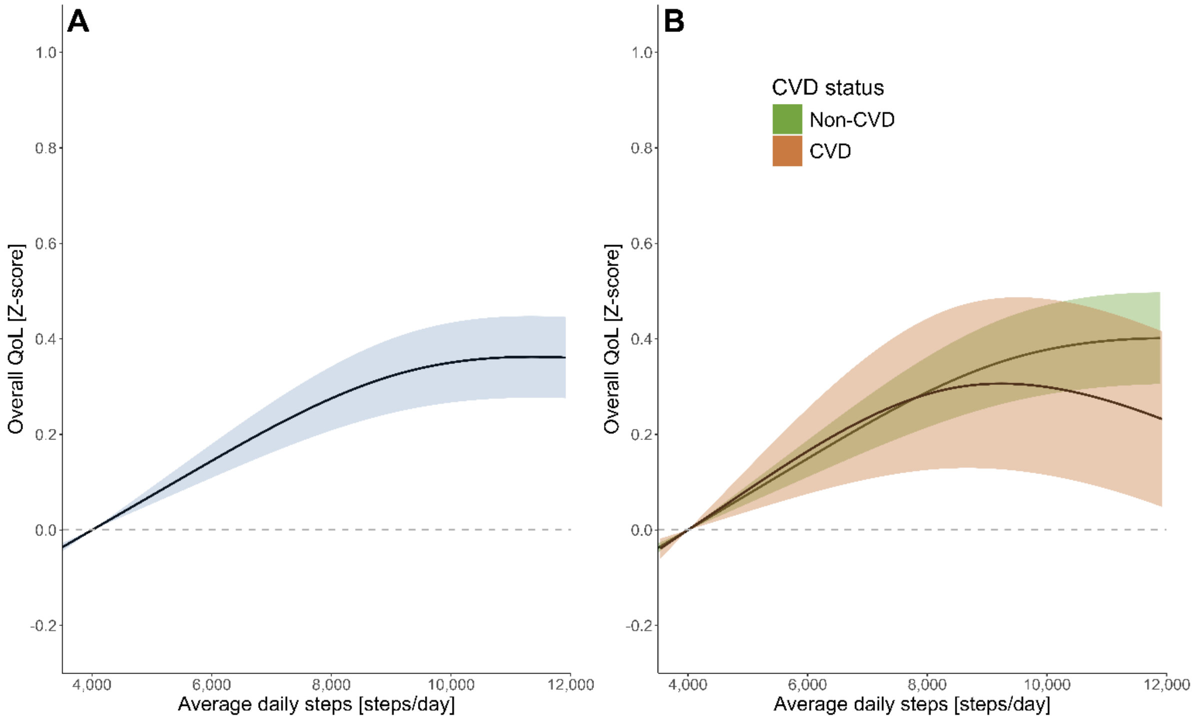

**Supplementary Figure 10. Dose-response association steps and overall quality of life.** The reference value is set at 4,000 steps/day. (A): overall association steps and quality of life excluding patients with chronic diseases. (B): overall association steps and quality of life excluding patients with chronic diseases (except CVD), stratified by CVD status. Individuals without specific CVD diagnosis were excluded from this analysis. In addition, we excluded individuals with a heart valve disorder or peripheral artery disease due to small sample sizes (n<90). Individuals with a heart rhythm disorder were excluded since the lower limit was higher than the reference of 4,000 steps/day. Shading represents the 95% confidence interval.
